# Regional and temporal variations affect the accuracy of variant-specific SARS-CoV-2 PCR assays

**DOI:** 10.1101/2021.11.08.21266083

**Authors:** Chamteut Oh, Palash Sashittal, Aijia Zhou, Leyi Wang, Mohammed El-Kebir, Thanh H. Nguyen

## Abstract

Monitoring the prevalence of SARS-CoV-2 variants is necessary to make informed public health decisions during the COVID-19 pandemic. PCR assays have received global attention, facilitating rapid understanding of variant dynamics because they are more accessible and scalable than genome sequencing. However, as PCR assays target only a few mutations, their accuracy could be compromised when these mutations are not exclusive to target variants. Here we show how to design variant-specific PCR assays with high sensitivity and specificity across different geographical regions by incorporating sequences deposited in the GISAID database. Furthermore, we demonstrate that several previously developed PCR assays have decreased accuracy outside their study areas. We introduce `PRIMES`, an algorithm that enables the design of reliable PCR assays, as demonstrated in our experiments to track dominant SARS-CoV-2 variants in local sewage samples. Our findings will contribute to improving PCR assays for SARS-CoV-2 variant surveillance.

**Importance:** Monitoring the introduction and prevalence of variants of concern (VOCs) and variants of interest (VOIs) in a community can help the local authorities make informed public health decisions. PCR assays can be designed to keep track of SARS-CoV-2 variants by measuring unique mutation markers that are exclusive to the target variants. However, the mutation markers can not be exclusive to the target variants depending on regional and temporal differences in variant dynamics. We introduce `PRIMES`, an algorithm that enables the design of reliable PCR assays for variant detection. Because PCR is more accessible, scalable, and robust to sewage samples over sequencing technology, our findings will contribute to improving global SARS-CoV-2 variant surveillance.

## Introduction

SARS-CoV-2 has had an unprecedented impact on public health globally. However, despite the availability of vaccines, emerging new variants, which may have better infectivity, transmissibility, and immune evasion, threaten global public health again (1, 2). Monitoring the introduction and prevalence of variants of concern (VOCs) and variants of interest (VOIs) in a community can help the local authorities make informed decisions regarding public health (3–5). In particular, wastewater-based epidemiology (WBE) has been applied across the globe to monitor SARS-CoV-2 circulating in a community (6–9). WBE could complement clinical diagnosis because WBE allows health authorities to monitor transmission levels in communities, including asymptomatic patients, without requiring excessive resources (10).

Although sequencing is considered the gold standard to identify SARS-CoV-2 lineages, PCR assays have attracted global attention for variant detection due to several advantages (11). First, PCR is a more accessible tool because the instruments and reagents are more affordable. Second, PCR is more scalable because it can analyze dozens or hundreds of samples in only a few hours, while sequencing takes a much longer time (>12 hours) (12). Third, PCR is more robust to sewage samples that have low concentrations of SARS-CoV-2 genomes and contain different types of impurities (13). These advantages are beneficial to ramp up capacity for SARS-CoV-2 surveillance and facilitate deployment in regions where there is a lack of access to sequencing facilities to initiate their variant monitoring systems.

PCR assays are composed of about 20 to 30 base pair long primers or probes designed to detect single or multiple loci that characterize a target variant. Importantly, PCR assays can only examine less than 100 bp long sequences, while sequencing produces reads that span longer genome regions (> 1000 bp). Meanwhile, each variant or sub-lineage of SARS-CoV-2 is defined by a group of different mutations located throughout the entire genome. Therefore, distinct variants may have the same mutations, reducing specificity when used in PCR assays (14).

In this study, we introduce a computational tool, `PRIMES` (PRIMer Efficacy Sleuth), that can be used to analyze sequences available in open source databases such as GISAID (15, 16), to predict the sensitivity and specificity of a PCR assay to detect specific pathogen lineages of interest (**Fig. 1a**). Moreover, for a given set of mutations characterizing the target variant, `PRIMES` can also identify a subset of variant-specific mutations for designing PCR assays with high specificity and sensitivity. Using `PRIMES`, we show multiple examples of previous PCR assays (13, 17, 18) that were successfully applied to certain study areas that might not work for other regions (**Fig. 1b**). We also demonstrate that the PCR assays designed using `PRIMES` successfully identify the dominant lineages in sewage samples from Champaign County, IL, USA. We conclude that PCR assays ought to be designed or modified considering regional and temporal variations and that *in silico* analyses using open source databases can address the weaknesses of PCR assays. These findings will allow PCR assays to be applied more reliably for SARS-CoV-2 surveillance.

**Fig. 1.**
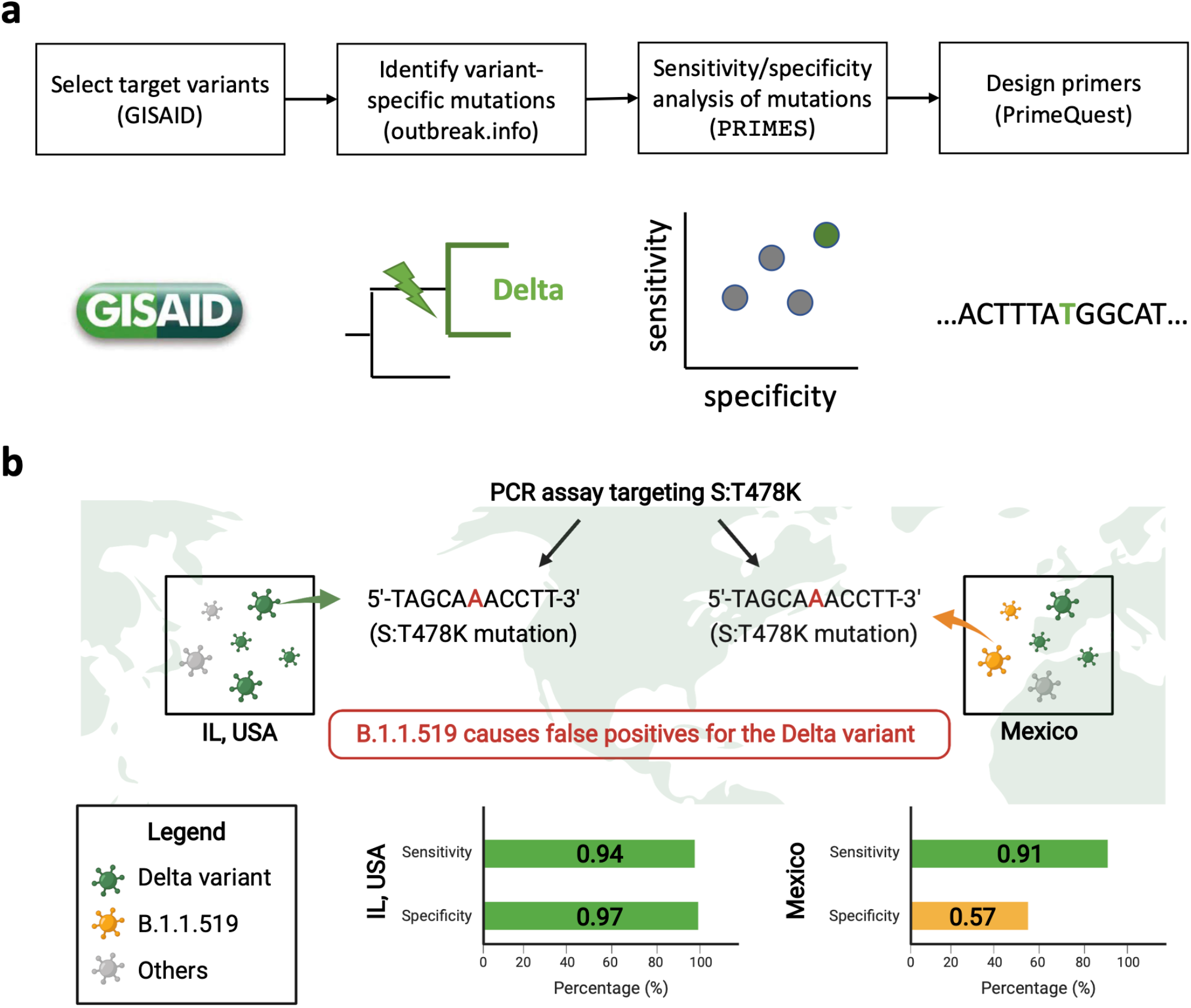
(**a**) A schematic describing the workflow for designing PCR assays while considering regional and temporal variations in GISAID samples. After selecting a target variant, we identify variant-specific mutations, which we rank in terms of sensitivity and specificity using the introduced tool PRIMES. Finally, we design primers for mutations with high sensitivity and specificity for the geographical region of interest. **(b)** An illustration showing the effect of regional lineages on the accuracy of variant-specific PCR assays.

## Results

### Analysis and design of PCR assays using PRIMES

As new variants of SARS-CoV-2 emerge and fade away throughout the world, a number of different lineages (1340 lineages as of August 2021) have been reported (19). Due to the evolutionary relationship of these lineages, they often share characteristic mutations. As such, PCR assays targeting only a few mutations (typically 1-3 mutations) have difficulty detecting samples from a specific lineage of interest with high specificity and sensitivity. In addition, while most lineages are limited to where they emerged, outbreaks of some lineages occasionally spread across the borders and become global concerns (such as variants of concern (VOCs) and variants of interest (VOIs)). Thus, regional and temporal differences in variant dynamics have to be considered for PCR assays.

The most widely used computational tool for assigning lineages to SARS-CoV-2 genomes is the Phylogenetic Assignment of Named Global Outbreak Lineages (Pangolin, https://pangolin.cog-uk.io/). Pangolin is a lineage designation pipeline that takes a FASTA file as input, containing one or more query sequences. Each query sequence is first aligned to the SARS-CoV-2 reference genome (Wuhan-Hu-1, NC_045512.2) using minimap2 v2.17 (20). After trimming the non-coding regions in the 5’ and 3’ ends of the aligned sequences, they are assigned to the most likely lineage out of all <u>currently designated lineages</u> using an underlying machine learning model referred to as PangoLEARN. The current version of PangoLEARN is a decision tree that is trained on data from GISAID that was manually curated with lineages.

By considering the lineage designation of Pangolin as ground truth, we perform an *in silico* analysis of the efficacy of PCR assays using `PRIMES` (available at https://github.com/elkebir-group/primes). Specifically, we search for the target sequence (containing a mutation targeted by the PCR assay) in each GISAID sequence and then estimate the overall specificity and sensitivity of the PCR assay. While this information is crucial in its own right, it can also be used to design lineage-specific PCR assays. Specifically, for a lineage of interest and a set of characteristic mutations, we use `PRIMES` to identify the set of mutations that should be targeted by PCR assays to get high specificity and sensitivity. We employ this approach to design PCR assays to detect the presence SARS-CoV-2 of both Alpha (e.g., B.1.1.7) and Delta (e.g., B.1.617.2) variants in sewage samples.

### Analysis of previously developed PCR assays

Here, we use `PRIMES` to analyze the efficacy of previously developed PCR assays targeting specific Spike (S) protein mutations in the SARS-CoV-2 genome. Recall that sensitivity and specificity are defined as follows.

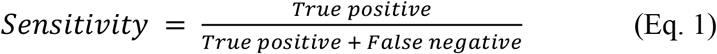

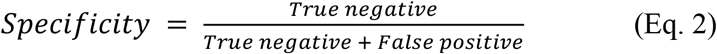

where true positive indicates the number of virus sequences that include the primer sequences and also belong to the lineage of interest. False negative is the number of virus sequences that do not include the primer sequences but belong to the lineage of interest. True negative is the number of virus sequences that do not include the primer sequences and do not belong to the lineage of interest as well. Finally, false positive indicates the number of virus sequences that include the primer sequences but belong to the lineage of interest. Strikingly, our analysis shows that several previously developed PCR assays would not be as accurate for samples collected from locations and periods beyond those included in the original study.

First, we focus on analyzing PCR assays targeting S:Δ69/70 to detect the Alpha variant (13, 17, 21). These PCR assays were verified with synthetic RNA controls and local sewage samples from Israel. Then, we simulated the application of these assays to sequences deposited in GISAID for Israel (n=13932 from January 2021 to October 2021). **Fig. 2a** shows that the Alpha variant was dominant (most prevalent variant) from January 2021 until May 2021, after which most samples were from other lineages. `PRIMES` predicts that the PCR assays targeting S:Δ69/70 (17) correctly assigned GISAID samples to the Alpha variants with a sensitivity of 0.95 and a specificity of 0.93. This finding can be attributed to the observation that the target mutations of these PCR assays, S:Δ69/70, is mostly exclusive to the Alpha variant in Israel, where this PCR assay was developed. However, although S:Δ69/70 was once a key mutation for the Alpha variant (B.1.1.7 first reported in February 2020), B.1.258.17 (first reported in August 2020) and B.1.620 (first reported in February 2021) and more lineages are also known to have the same mutation. Although these SARS-CoV-2 lineages are not significant in Israel (29/13932), they have significant prevalence in certain regions at some points in time due to local outbreaks. For example, the B.1.258.17 lineage accounted for 21.8% of all the sequences on GISAID from Slovenia for the period between January 2021 to October 2021.

**Fig. 2.**
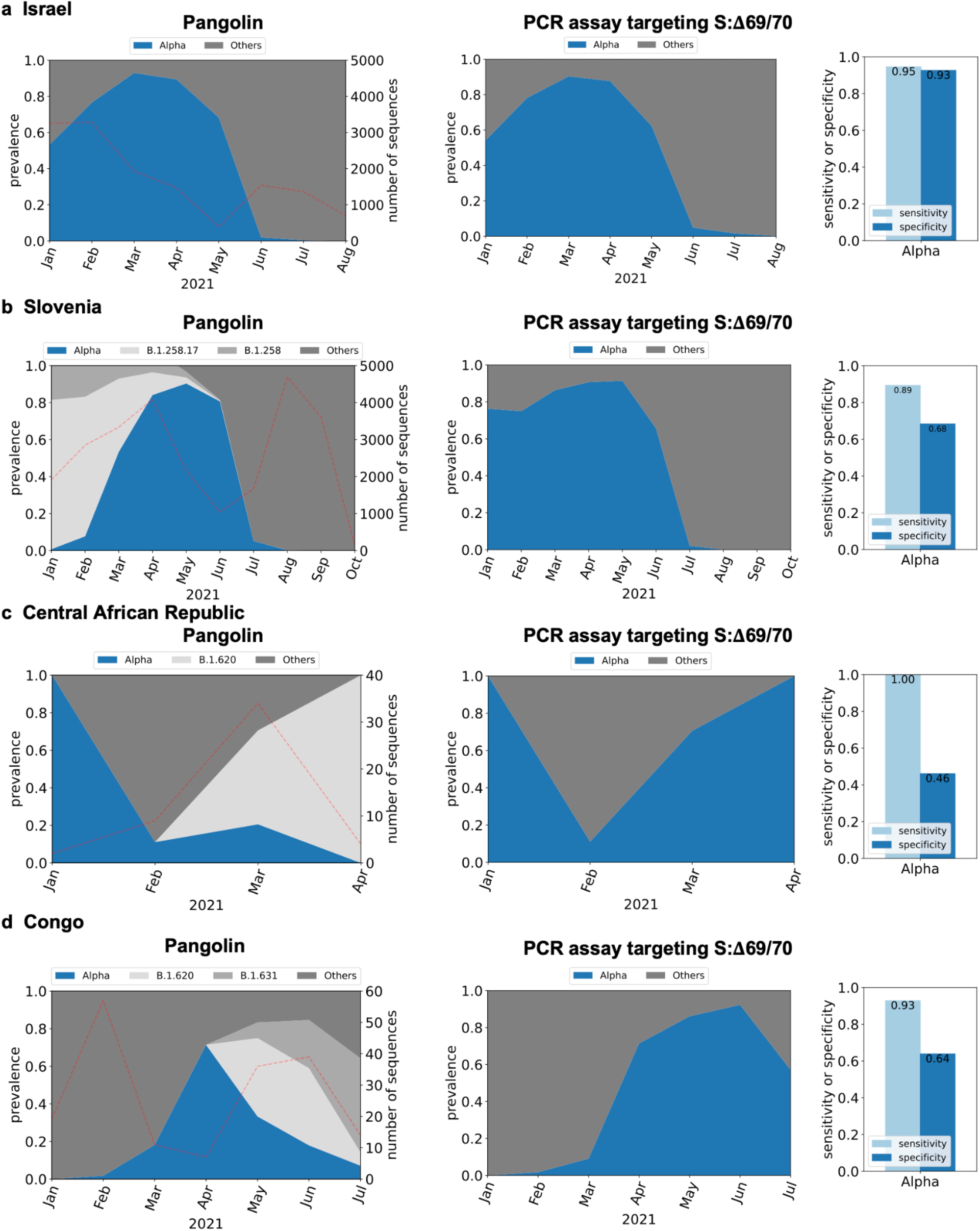
*In silico* analysis of PCR assay targeting S:Δ69/70 mutation (17) to detect the Alpha variant for GISAID samples from (**a**) Israel (n=13,932), (**b**) Slovenia (n=25,528), (**c**) Central African Republic (n=49) and (**d**) Congo (n=183). Dotted lines on left figures indicate the number of sequences used for the *in silico* analyses.

While this PCR assay targeting S:Δ69/70 was only applied to wastewater samples from Israel in the previous study (17), we can use `PRIMES` to predict the sensitivity and specificity of this PCR assay on GISAID samples from any other region. **Fig. 2b** shows our analysis on samples from Slovenia (n=25528, from January 2021 to October 2021) where the prevalence of lineage B.1.258.17 was significant until May 2021 and dominating in January 2021 and February 2021. On the other hand, Alpha variant has less than 10% prevalence in January 2021 and February 2021. However, our analysis of the PCR assays shows that the Alpha would have been dominant from January 2021 until June 2021. This error came from the fact that these assays would have incorrectly assigned genomes belonging to the B.1.258 and B.1.258.17 lineages to the Alpha variant. The false positives for the Alpha variant continued until June 2021 the B.1.258.17 lineage fades out. Thus, while the estimated sensitivity of the assay for the Alpha variant in Slovenia is 0.89, the specificity is estimated to be only 0.68. We also found that PCR assays targeting S:Δ69/70 could lead to significant amounts of false positives when applied to samples from the Central African Republic (**Fig. 2c** shows that the estimated specificity is only 0.46 due to samples from B.1.620), Republic of Congo (**Fig. 2d** shows that the estimated specificity is only 0.64 due to B.1.620 and B.1.631).

We conducted a similar analysis for another PCR assay targeting mutation S:Δ144/145 to detect the Alpha variant (13). This assay was applied to samples from wastewater treatment plants and selected residential buildings across the USA to track the occurrence of the Alpha variant over time in 19 communities. **SI Fig. 1a** shows that this assay works well for GISAID sequences from the USA with estimated sensitivity and specificity of 0.90 and 0.98, respectively. However, several lineages including C.1.2, B.1.620, B.1.1.318, B.1.525 (or the Eta variant), B.1.637, B.1.625, and AZ.2 also have the same mutation S:Δ144/145 targeted by this assay. This PCR assay can produce false results if any of the aforementioned lineages have significant prevalence in the area being studied. For example, we analyzed GISAID sequences collected from Gabon (n=254, from January 2021 to May 2021). **SI Fig. 1b** suggests that Gabon had significant sequences from the Eta variant and the B.1.1.318 lineage from February 2021 to May 2021, both of which have the target mutation S:Δ144/145 mutation. As a result, even though the number of sequences from the Alpha variant increased from February 2021 to April 2021 and then decreased in May 2021, the PCR assay would have predicted a continuous increase in the prevalence of the Alpha variant from February 2021 to May 2021 (**SI Fig. 1b**). The estimated specificity of this assay for detecting the Alpha variant on GISAID sequences from Gabon is only 0.74. We see a similar result by analyzing GISAID sequences from Togo (n=157 from January 2021 to April 2021) in **SI Fig. 1c**. In fact, many countries in Africa, including Nigeria and Ghana, were also expected to have lower specificity for the PCR assay targeting S:Δ144/145 because of B.1.1.338 and B.1.525 lineages (**SI Table 1**).

This propensity for false positive results is not limited to PCR assays developed to detect samples from the Alpha variant. We demonstrate this by considering a recent PCR assay targeting mutation S:T478K of the Delta and Delta plus lineages (18). However, this mutation is also present in the B.1.1.519 lineage. This lineage accounted for only around 1.2% of sequences from the USA (n=1,187,412 from January 2021 to October 2021), so the PCR assay targeting S:T478K was expected to work well for IL, USA, showing 0.94 of estimated sensitivity and 0.97 of estimated specificity (**Fig. 3a**). However, the B.1.1.519 lineage was dominant in Mexico (n=28,956) and explained 30% of the total GISAID sequences from January 2021 to October 2021 (**Fig. 3b**). Therefore, our analysis shows that the PCR assay targeting S:T478K would estimate that the Delta variant was dominant all the way from January 2021 to October 2021, when in reality, Delta variant sequences were collected and later deposited in GISAID, starting May 2021 (**Fig. 3b**).

**Fig. 3.**
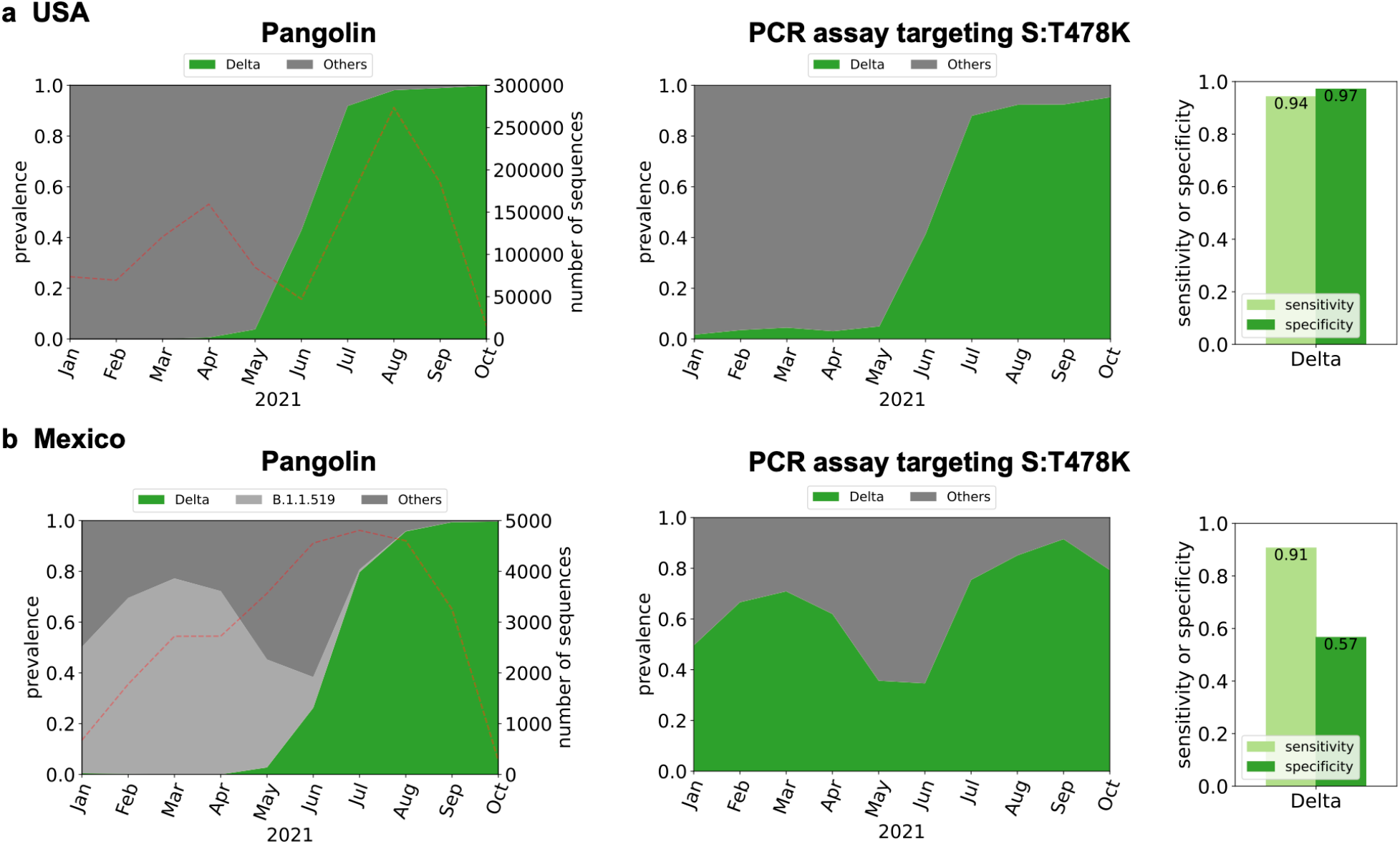
*In silico* analysis of PCR assay targeting S:T478K mutation (18) to detect the Delta variant: (**a**) USA (n=1,187,412) and (**b**) Mexico (n=28,956). Dotted lines on left figures indicate the number of sequences used for the *in silico* analyses.

The examples of regional and temporal characteristics affecting the accuracy of PCR assays for the detection of SARS-CoV-2 samples of specific lineages of interest are not limited to the cases mentioned above. Globally, only a few VOCs and VOIs accounted for higher than 1% of the total sequences in GISAID, while most of other SARS-CoV-2 lineages explain less than 1%. However, as we examine narrower regions, we may find outbreaks of certain lineages that could be overlooked when we focus on the prevalence on a global scale. For example, as of October 2021, B.1.526 lineage accounted for 1% of reported sequences in the world. However, the prevalence of the lineage increases as we narrow down the study area to local: 4% in the U.S., 17% in New York state, and 30% in Bronx country. In the case of B.1.429 lineage, the prevalences are 1%, 4%, 11%, and 38% in the world, the U.S., California State, and Riverside County, respectively. The number of B.1.258 lineage was less than 0.5% of total sequences worldwide, but it accounted for 54% of cases in Cyprus.

We summarized SARS-CoV-2 lineages that have the same mutation that our target variant possesses in **SI Table 1**. When we use PCR assays targeting certain mutations, this table will help identify the lineages that would interfere with our PCR assay. In **SI Table 2**, we also tabulated countries where each of the SARS-CoV-2 lineages summarized in **SI Table 1** accounted for higher than 1% of total sequences. This table will explain whether the lineages that would interfere with your PCR assays are dominant in the study areas. By interpreting **SI Table 1** and **SI Table 2** together, we can find various examples where certain PCR assays would not work reliably. In conclusion, the findings that previously developed PCR assays would not work for certain regions due to the presence of lineages sharing some unique mutations with other lineages motivated us to establish PCR assays for variant detection based on the characteristics of sequences reported from our target study area (IL, USA).

### Design of variant-specific PCR assays considering regional and temporal characteristics

We describe the proposed workflow to design variant-specific PCR assays considering regional and temporal variant dynamics using `PRIMES` (**Fig. 1a**). Our goal is to design variant-specific PCR assays to track variants with significant prevalence in the USA, with a particular focus on the state of Illinois.

First, we investigated the prevalence of SARS-CoV-2 lineages in our regions of interest to select lineages that we need to track. To this end, we downloaded 1,187,412 SARS-CoV-2 sequences from GISAID collected between January 2021 and October 2021 in the USA, including 20,165 sequences collected in Illinois. These sequences were assigned to the most likely lineage using Pangolin (**Fig. 4a, b**). Focusing on the variant dynamics in the state of Illinois (**Fig. 4a**), we observe that the B.1.2 lineage was dominant from January (61%) to February (44%), eventually giving way to the Alpha variant. The Alpha variant became dominant in April (44%), May (61%), and June (62%). Then, the Delta variant samples replaced the Alpha variant samples and has been the dominant lineage in the state (95% in July and >99% in August, September and October). Other VOIs and VOCs, including Epsilon, Iota, and Beta variants, accounted only for 2.2%, 1.5%, and 0.4% of total sequences, respectively. Similar trends were observed in sequences collected throughout the USA (**Fig. 4b**). Based on the variant dynamics of our regions of interest, we decided to design PCR assays to enable monitoring of the two major variants, the Alpha and the Delta variants.

**Fig. 4.**
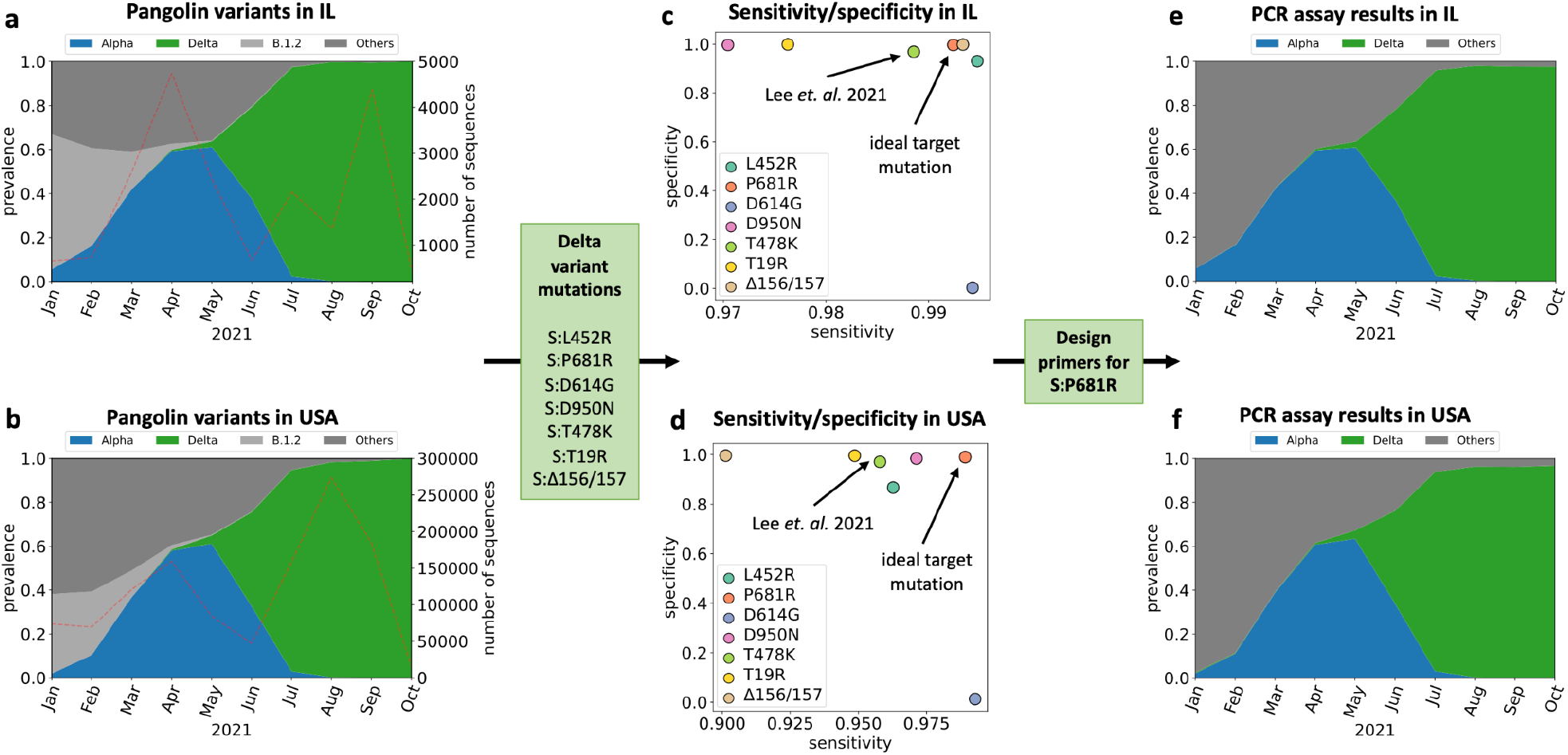
Variant dynamics determined by Pangolin using GISAID samples from **(a)** the state of Illinois in USA (n=20,165) and (**b**) USA (n=1,187,412). Focusing on the spike protein mutations in the Delta variant, we show the sensitivity and specificity of assigning the Delta variant based on the presence of each mutation in GISAID samples from **(c)** IL and **(d)** USA. The estimated assignment of GISAID samples from **(e)** IL and **(f)** USA to variants using the primer designed to target mutation S:P618R.

Second, we designed PCR assays to find unique mutations that are exclusive to our lineage of interest. We utilized https://covariants.org to list up nonsynonymous mutations that define target variants (**SI Table 1**). We focused on mutations located in the spike gene, which has a higher frequency of mutation than other SARS-CoV-2 genes (22). Previous studies have shown that primers targeting mutations in the spike gene enable accurate detection of SARS-CoV-2 lineages in sewage samples with low virus concentration (21). As a result, for the Alpha variant, we identify 9 mutations in the spike gene – S:Δ69/70, S:Δ144, S:N510Y, S:A570D, S:D614G, S:P681H, S:T716I, S:S982A, and S:D1118H. For the Delta variant we identify 7 mutations – S:T19R, S:Δ156/157, S:L452R, S:T484K, S:D614G, S:P681R, and S:D950N. Note that mutation S:T484K was targeted by the PCR assay to detect the Delta and Delta plus variant (18).

Third, we used `PRIMES` to compute the sensitivity and specificity of lineage assignments performed using each of the selected mutations. We assumed if the specificity and sensitivity of the mutations are higher than 0.99, the mutations are exclusive to the target variant. This criterion allows us to identify the ideal target mutation for the design of the PCR assay that would yield high specificity and sensitivity in our regions of interest. For the Alpha variant, we found three acceptable mutations (S:A570D, S:T716I, and S:S982A) (**SI Fig. 2b**), and we chose the S:A570D mutation because PCR assay targeting S:A570D has already been verified to work for sewage samples, we adopted this mutation in our analysis (13). For the Delta variant, we found three acceptable mutations (S:L452R, S:P681R, and S:Δ156/157) with GISAID samples from the state of Illinois. However, if we look at all GISAID samples from the USA, the sensitivity for S:L452R and S:Δ156/157 mutations to characterize the Delta variant drops below 0.97. Thus, regional variation can lead to a drastic change in the performance of variant-specific PCR assays. Since our goal is to develop PCR assays that are effective in other states besides Illinois in the USA as well, we instead choose S:P681R, which has high sensitivity and specificity in both Illinois (sensitivity is 0.99 and specificity is 0.99) as well as the USA (sensitivity is 0.99 and specificity is 0.99). Importantly, this mutation has higher sensitivity and specificity in both regions of interest compared to mutation S:T478K (sensitivity is 0.99 and specificity is 0.97 in Illinois, while sensitivity is only 0.96 and specificity is only 0.98 in the USA, see **Fig. 4c,d**) that previously targeted to monitor the Delta and Delta plus variants (18).

The fourth step is to design the allele-specific primers for the selected mutations. Since both our selected target mutations are single nucleotide polymorphisms (SNPs), we designed allele-specific qPCR assays in which either a forward or a reverse primer target the SNP at the 3’ end with a mismatch near the SNP location to improve the specificity of the assays (13). All RT-qPCR assays were designed using PrimerQuest (Integrated DNA Technologies; IDT, USA) to have an annealing temperature from 58 to 63 °C for primers and GC contents from 30 to 60%.

Finally, we can estimate the efficacy of the candidate RT-qPCR assays using `PRIMES`. Specifically, we determine the sensitivity and specificity of our assays on the GISAID samples collected from the regions of interest by searching for sequences of a forward primer and a reverse primer in each query sequence. Note that the sequences of reverse primers were converted to reverse sequences to have all sequences, including primers and viruses, on the same strand. If the viral sequence includes the forward and reverse sequences, we assumed that the PCR assay would detect the viral sequence (an illustrative example is shown in **SI Fig. 3**). We note that operational failures of the assay due to inappropriate primer design or PCR inhibitors are not considered by `PRIMES`. Some lineages should be expected to lower the sensitivity or specificity of our assays based on **SI Table 1** (e.g., B.1.1.189, C38, and B.1.636 for the Alpha variant detection and AU.3, AU.2, P.1.8, B.1.617.3, A.23.1, B.1.617.1, B.1.551, B.1.466.2, B.1.1.528, Q.4, B.1.623, B.1.1.25, C.36, and AY.28 for the Delta variant detection), but importantly those lineages were not detected or had very low prevalence in our regions of interest. The estimated sensitivity and specificity for PCR assays designed to detect viruses from the Alpha and Delta variants were all high for our study scope, and in Illinois in particular (sensitivity is 0.99 and specificity is 0.99 for detection of Alpha variant and sensitivity is 0.98 and specificity is 1.00 for detection of the Delta variant in Illinois, see **SI Fig. 4**, for sensitivity and specificity of detecting the two variants in GISAID samples from all of the USA). These values are higher than the sensitivity and specificity estimated for the previously developed PCR assays in their regions of interest (see **Fig. 2** and **3**). In the subsequent section, we demonstrate this performance of our PCR assays translated to synthetic controls and actual sewage samples collected in our community.

### Verification of PRIMES-designed PCR assays by synthetic RNA controls

We applied the RT-qPCR assays designed with `PRIMES` to synthetic RNA controls for WT, Alpha, and Delta variants to experimentally confirm the sensitivity (i.e., the limit of quantification; LOQ and limit of detection; LOD) and specificity (i.e., cross-reactivity). Regarding sensitivity, we found that the LOQs for total SARS-CoV-2, Alpha variant, and Delta variant were all 10 gene copies (gc)/µL or 50, 30, and 30 gc/reaction, respectively (**SI Fig. 5a**). Also, the LODs of RT-qPCR assays for total SARS-CoV-2, Alpha variant, and Delta variant were 1.0, 1.3, and 1.3 gc/µL or 5.0, 3.9, and 3.8 gc/reaction, respectively (**SI Fig. 5b**). Because LODs for our assays were close to the theoretical LODs of RT-qPCR (3.0 gc/reaction)(23), we concluded that our RT-qPCR assays are sensitive to detect RNA of target variants.

As for the cross-reactivity, we found that when the concentrations of the RNA synthetic control were high (i.e., 10^4^ and 10^5^ gc/µL), we detected Cq values from WT RNA controls. This finding suggests that the presence of WT caused false positives for the Alpha variant detection (**SI Fig. 6a**). However, the Cq value differences between the Alpha variant and WT were greater than 11 that is about 10^3^-fold difference in RNA concentrations. This difference in Cq values is equivalent to less than 0.1% error when quantifying the Alpha variant, and thus we considered this error is acceptable for our study. When the RNA synthetic control concentrations were low (i.e., less than 10^3^ gc/µL), the Cq values from WT were lower than LOD, which will be disregarded in this study, so the false positives were not detected. We found similar results from the specificity experiments for the Delta variant assay (**SI Fig. 6b**). When the concentrations of the RNA synthetic controls were high (i.e., 10^4^ and 10^5^ gc/µL), the Cq value differences from Delta variant and WT were greater than 13. At the lower concentration (i.e., less than 10^3^ gc/µL), the Cq values from WT (i.e., false positives) were less than LOD. Because the measured cross-reactivities by WT were negligible, we concluded our RT-qPCR assays are specific to measure target variants.

We further confirmed the applicability of `PRIMES`-designed PCR assay to determine predominant variants in mixtures of synthetic RNA controls. The results from the mixtures of synthetic RNA controls are presented in **Fig. 5a** and **5b**. The y-axis shows the prevalences, which are the ratios of each variant’s concentration to total SARS-CoV-2 concentration. The variant showing the highest prevalence became the dominant variant. If none of the two targets (i.e., Alpha and Delta variants) are higher than 0.5, the Others, which indicate all SARS-CoV-2 lineages other than our target variants (i.e., the Alpha and Delta variant), became the dominant variant. With the highest total virus concentrations (10^4^ gc/µL), our RT-qPCR assays successfully assign the correct dominant variant to all experimental cases (p < 0.001) (**Fig. 5a**). For example, in the case of the mixtures between WT and Alpha variant, we assigned Alpha variant to the RNA mixtures whose actual prevalence of Alpha variant were 0.7 and 0.9 whereas we assigned ‘Others’ when the prevalence of Alpha variant were 0.1 and 0.3. In the same manner, we also assigned the dominant variant correctly to the mixtures of Alpha and Delta variants. Specifically, we assigned the Alpha variant when the actual prevalences of Alpha variant were 0.7 and 0.9. At the same time, the Delta variant was assigned to the other two mixtures whose prevalences of Alpha variant were 0.1 and 0.3. In addition, we found that the PCR assays assigned the dominant variant correctly when the total virus concentrations were 10^1^ gc/µL for all mixing ratios (**Fig. 5b**). However, the statistical analysis showed that the comparisons of prevalences determined by the RT-qPCR were significant only when the mixing ratios were 0.9:0.1 or vice versa for mixtures of WT and Alpha variant or Alpha variant and Delta variant. Note that when total virus concentrations were 10^1^ gc/µL, concentrations of each synthetic RNA control were ranging from 1×10^0^ to 9×10^0^ gc/µL depending on the mixing ratios, which were less than their LOQs (10^1^ gc/µL). Based on these findings, we concluded that our RT-qPCR assays could find the dominant variant when total SARS-CoV-2 concentrations are higher than LOQs (10^1^ gc/µL) and the prevalence of target variants is higher than 0.9 (**Fig. 5b**). When the total SARS-CoV-2 concentrations become higher, Alpha and Delta variant concentrations are higher than LOQs of the RT-qPCR assays (10^1^ gc/µL), our RT-qPCR assays can assign the dominant variant when its prevalence is higher than 0.7 (**Fig. 5a**).

**Fig. 5.**
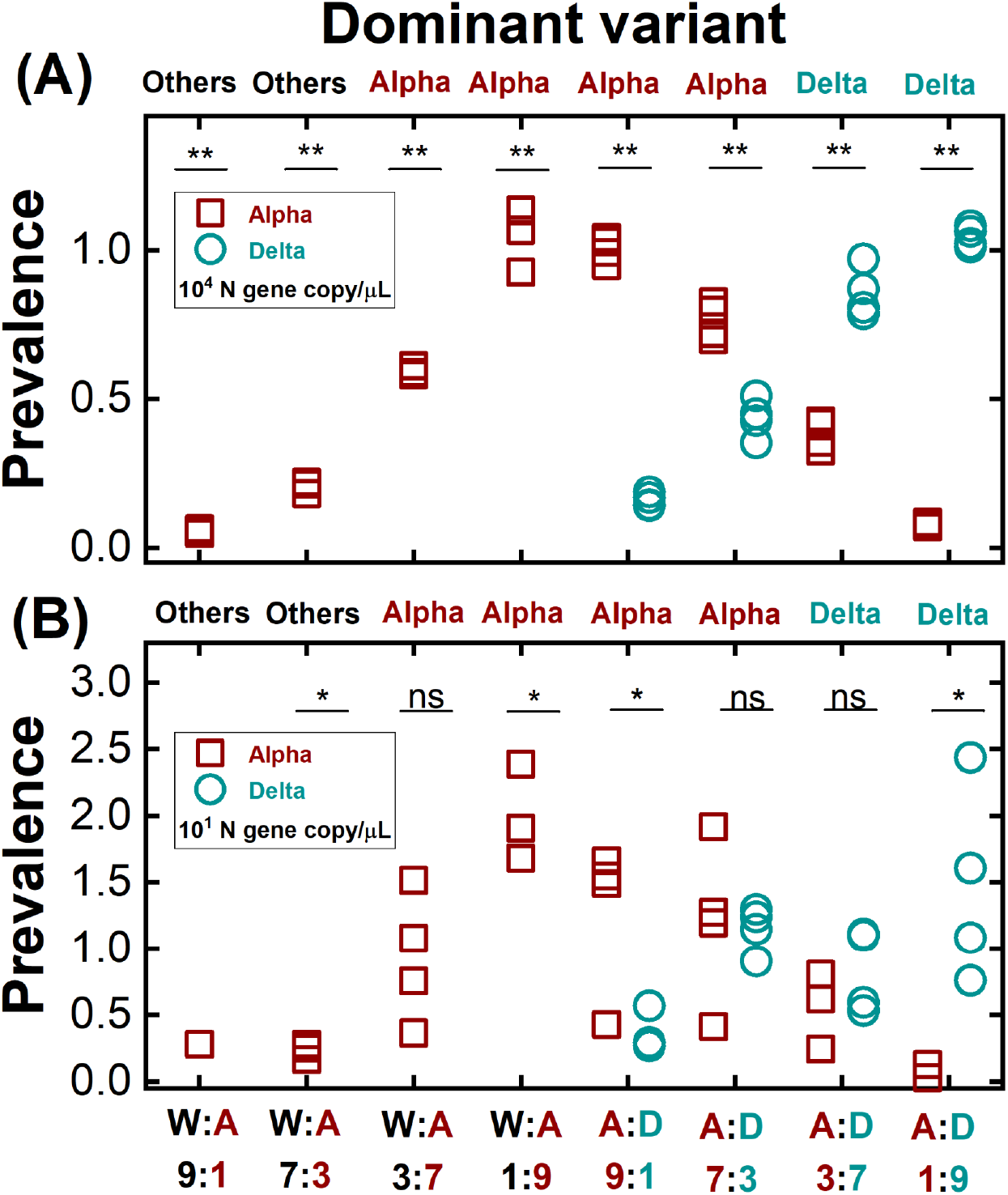
The dominant variants of the mixtures of synthetic RNA controls determined by RT-qPCR assays. Total SARS-CoV-2 concentrations were determined by the N gene concentrations at **(a)** 10^4^ gc/µL and **(b)** 10^1^ gc/µL. Prevalences on the y-axis indicate the ratio of each variant’ concentration to the total virus concentrations. The x-axis presented the mixing ratios of different synthetic RNA controls (W, A, and D stand for WT, Alpha variant and Delta variant, respectively). Decisions made on top of each graph showed the dominant variant determined by the RT-qPCR assays. One sample t-test or two-sample t-test were conducted to compare prevalence between Alpha variant and 0.5 or the prevalences between Alpha and Delta variants (ns: p>0.05, *: 0.001<p<0.05, **:p<0.001), respectively.

### Application of PCR assays to sewage samples and confirmation by NGS

We applied our PCR assays to six different local sewage samples. We first obtained RNA extracts from those sewage samples. The total SARS-CoV-2 concentrations (i.e., N gene) of these RNA extracts ranged from 1.4 × 10^*1*^ to 1.8 × 10^*2*^ gc/µL (**SI Table 3**). After accounting for recovery efficiencies and concentration factors, the SARS-CoV-2 concentrations (i.e., N gene) of these sewage samples ranged from 1.3 × 10^*3*^ to 6.0 × 10^*4*^ gc/L (**Eq. 3**). These concentrations agree with the SARS-CoV-2 concentrations of sewage samples analyzed previously (24). Then, we determined the prevalence of variants based on the ratios of the Alpha variant concentration (determined by `PRIMES`-designed PCR) over the total SARS-CoV-2 (N gene). We found that sample #1 has 0.85 prevalence of the Alpha variant. Based on the results with the RNA synthetic mixtures, we assigned the Alpha variant as a dominant variant to sample #1. Similarly, we assigned the Delta variant to Sample #5 and #6 because of their prevalences of 0.92 and 0.73, respectively. On the other hand, none of the Alpha and Delta variants presented higher than 0.5 prevalence for Sample #2, #3, and #4, so we assigned Others to these three samples.

To further confirm whether the RT-qPCR results were correct, we conducted NGS analysis to examine eight mutation markers for the Alpha variant (S:Δ69/70, S:Δ144, S:N501Y, S:A570D, S:P681H, S:T716I, S:S982A, and S:D1118H) and six mutation markers for the Delta variant (S:T19R, S:Δ156/157, S:L452R, S:T478K, S:P681R, and S:D950N) on the spike gene of two sewage samples (#5 and 6) and three synthetic RNA controls (WT, Alpha, and Delta variant). Samples #1, #2, #3, and #4 were not appropriate for sequencing due to the low SARS-CoV-2 concentrations (<10^2^ gc/µL). From Sample #5 and #6 classified to the Delta variant by the RT-qPCR assays, we detected all six mutations for the Delta variant. In comparison, none of the eight mutations for the Alpha variant were detected in these samples. We believe the NGS analyses were reliable because of the results with the synthetic RNA controls. We detected all mutation markers with corresponding synthetic RNA controls. For example, we detected the eight Alpha variant mutations from the Alpha variant RNA samples and found the six Delta variant mutations from Delta variant RNA controls (**Table 1**). Therefore, the agreement between the NGS analysis and RT-qPCR assays supports that our RT-qPCR can assign the most likely variant for the local sewage samples.

**Table 1.**
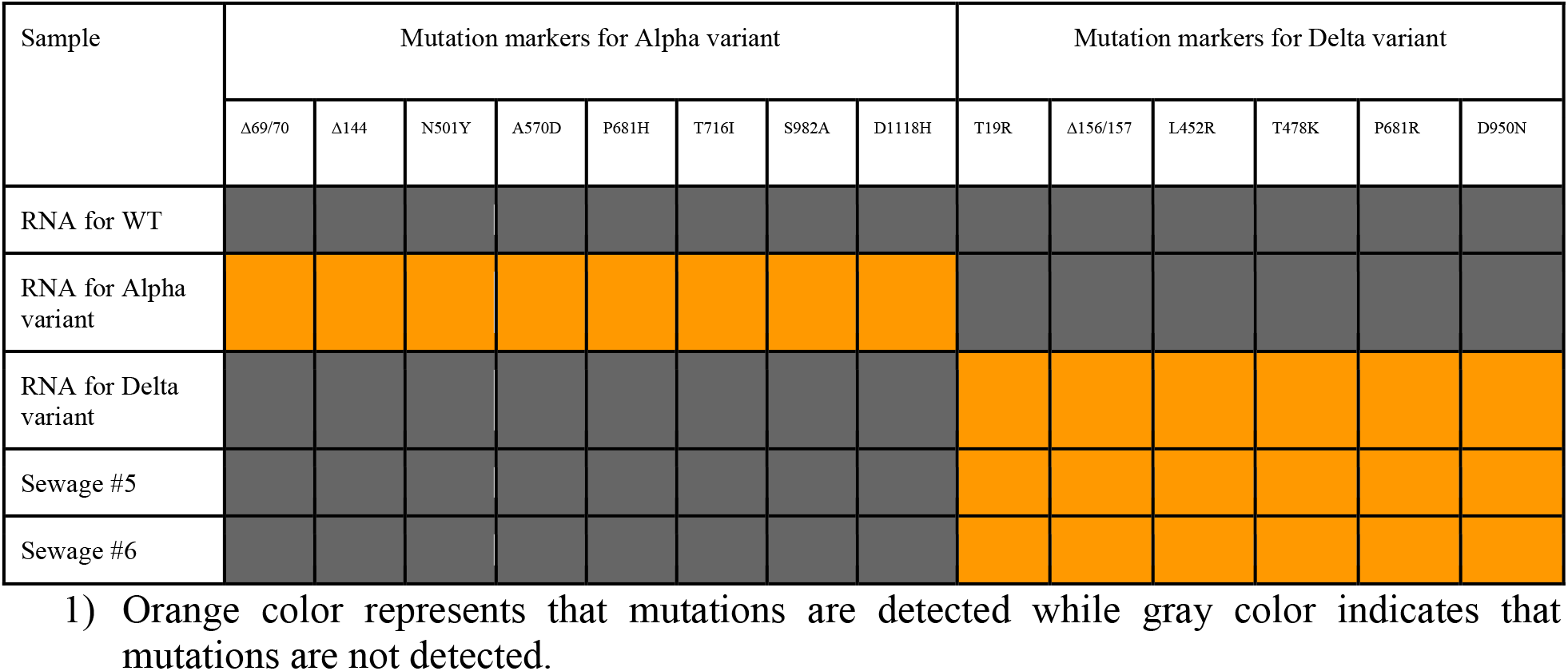
Comparisons between RT-qPCR assays and NGS analysis

## Discussion

PCR assays have advantages for SARS-CoV-2 variant detection in sewage over sequencing technologies such as low cost, fast turnaround, and robustness to environmental samples. However, PCR assays can examine only a few mutations due to size constraints of primer and probe sequences, compromising their accuracy as distinct SARS-CoV-2 lineages may share target mutations. We used the `PRIMES` algorithm to show that the current variant-specific PCR assays have diminished accuracy when applied outside the region where they were developed. These findings suggest that considering regional and temporal dynamics of variants is important to secure the sensitivity and specificity of PCR assays that target only a limited number of mutations. Subsequently, we used `PRIMES` and open-source databases (e.g., GISAID, Pangolin, or outbreak.info) to design PCR primers to determine the dominant SARS-CoV-2 variants (i.e., Alpha and Delta variant) in local sewage samples.

The regional and temporal variations are especially critical for SARS-CoV-2 detection because various SARS-CoV-2 lineages with different genotypes have been reported across the world. Commercial PCR kits for variant detection are currently available. However, these kits also target a few mutation markers originating from SARS-CoV-2 lineages of interest (25–27). As we showed above, targeting a single mutation could make the assay less accurate at certain regions due to the presence of other lineages that have the same mutation. In addition, our findings are not limited to PCR assays, but are also relevant for the other types of molecular assays such as loop-mediated isothermal amplification (LAMP), PfAgo-based assay, and Clustered, Regularly-Interspaced Short Palindromic Repeats (CRISPR)-based assays that are designed to detect specific RNA sequences for virus detection (28–30). For example, two LAMP assays that target N genes for SARS-CoV-2 test could have low accuracy when applied outside of Germany or the US, where the assays were developed and verified with clinical samples (28, 31). This low accuracy issue could happen because their primers include sequences for <u>N:A119S</u>, a mutation marker for the Zeta variant (P.2 lineage). The Zeta variant was dominant in some South American countries (Suriname, Paraguay, Uruguay, and Brazil). Zhang et al. (2021) (32) designed a LAMP assay targeting the ORF1a gene, but their F2 primer includes sequences for ORF1a:V86F and F1 primer ORF1:E102K. The Food and Drug Administration (FDA) also recommended that mutations present in the sequences where molecular diagnostic tests target for virus detection should be monitored by *in silico* analysis (33). Our `PRIMES` tool allows users and developers of molecular diagnostic assays to follow this recommendation.

Perfect loci to target viral mutation are not realistic because viruses evolve randomly, so one that looks perfect could be affected by emerging variants. For example, the S:Δ69/70 mutation used to be a unique mutation for the Alpha variant, but the Eta variant, which appeared later, also has the same mutation. Thus, if the S:Δ69/70 is considered an exclusive mutation to the Alpha variant, the Eta variant will be false positive for the Alpha variant. In addition, sub-lineages in the target variant may not have one of the mutation markers for the target variant. For example, less than 0.5% of Q.4 (one of the sub-lineages for the Alpha variant and reported December 2020) is known to have a S:P681H. The S:P681H mutation is one of the mutation markers for the Alpha variant. Thus, if S:P681H mutation is targeted for the Alpha variant, Q.4 will cause false negatives. These examples demonstrate that PCR assays could have different sensitivities and specificities depending on various lineages of SARS-CoV-2 that coexist with the target lineage.

Global genomic databases for emergent variants have greatly improved since the onset of COVID-19 pandemics (34). Before COVID-19, influenza sequences are archived in GISAID. Fast mutating pathogens such as influenza and coronavirus should be monitored because they have pandemic potential. As we showed in this study, assays targeting these pathogens need to keep up with their evolution, and the developed methodology facilitates genomic surveillance of any fast mutating pathogen.

## Methods

### Sewage samples

We followed the Minimum information Environmental Microbiology Minimum Information (MIQE) Guidelines to ensure the credibility and reproducibility of our data (35). Detailed information on the MIQE is summarized in **SI Table 4**. Also, detailed information from sample collection to data analysis is described in **SI Table 5**. Briefly, we used ISCO automatic samplers (6712 ISCO, Teledyne ISCO, USA) to collect three-day composite sewages (about 2 L) from the sewer distribution system across Champaign County, IL, USA. MgCl2 was added to the sewage samples at the final concentration of 50 mM to facilitate the coagulation of viruses and sewage sludge. We kept sewage samples on ice while moving them to our laboratory in 2 hours. We gently removed the supernatant upon arrival and added 200 μL of bovine coronavirus (BCoV) to the remaining solution (about 50 mL) to determine virus recovery efficiency. After 5 minutes of incubation at room temperature, we centrifuged the mixture at 10,000 rpm (13,900 g) for 30 minutes (Sorvall Legend RT Plus, Thermo Fisher Scientific, USA). The supernatant was discarded again, and the sludge (about 1 g) was taken to harvest viruses. Then, we extracted viral RNA from the sludge using the Viral RNA extraction Mini Kit (Qiagen, Germany) following the manufacturer’s procedure. The RNA extracts were purified using an RNA purification kit (RNeasy MinElute Cleanup Kit, Qiagen, German) to reduce the PCR inhibitions. It took less than 9 hours from sample collection to RNA extraction. The RNA samples were stored at -80°C until downstream analyses were ready. The same sample preparation processes were applied to drainages discharged from a food processing industry whenever we processed sewage samples. There are no sources of human feces that merged to these drainages, which were therefore used for negative controls. Indeed, we did not detect any SARS-CoV-2 from these negative controls. Therefore, we are confident there were no false positives for SARS-CoV-2 in our sewage samples. With the concentrations of RNA extracts, we used Eqs. 3-5 to determine the virus concentrations in sewage samples.

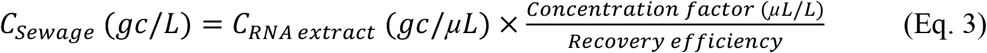

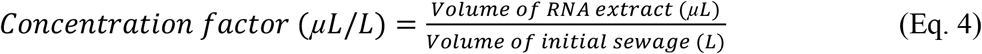

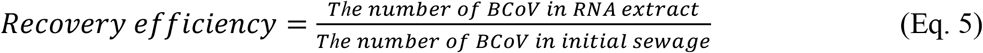

### Determination of LODs and LOQs

We first determined the limit of detection (LOD) and the limit of quantification (LOQ) for Alpha and Delta variants with serial dilutions of the synthetic RNA controls. We prepared 10-fold serial dilutions of synthetic RNA controls and determined a positive sample fraction at each concentration. The number of replicates for concentrations near LOD are 20 while the sample numbers for the higher concentration were 4. We used a sigmoidal function (**Eq. 6)** to determine the trend lines for fraction positive samples with different concentrations and calculated the LODs (36).

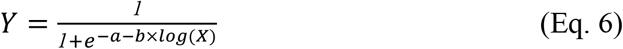

where X is gene copy (gc/μL), Y means positive rate, and both a and b are constants. LOQ was defined as the lowest concentration with coefficient of variation (CV) less than 35% (36). We calculated CV by **Eq. 7**.

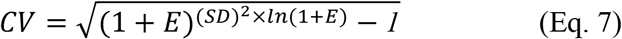

where E is a qPCR efficiency, SD is a standard deviation of Cq values.

### PCR assays for SARS-CoV-2 variant detection in synthetic RNA control

We applied the RT-qPCR assays to 10-fold serial dilutions of synthetic RNA controls to determine LOQs and LODs. LOQ was defined as the lowest concentration with coefficient of variation (CV) less than 35% (36) and LOD was defined as the concentration at which RNA samples test positive (i.e., Cq<40) with 95% probability. For example, we applied the RT-qPCR assay for Alpha variant to 10-fold serial dilutions of Alpha variant RNA controls and those of the WT RNA controls, then compared the Cq values from Alpha variant and WT.

We applied each of RT-qPCR assays for Alpha and Delta variant to the synthetic controls of its target variant and WT to determine the cross-reactivity. This process is important because our assays for the Alpha and Delta variants were designed to detect only a SNP of target variants among other lineages that do not have the same SNP. In this experiment, we mixed synthetic RNA controls of WT and Alpha variant or Alpha and Delta variant because these two mixtures represented the transitions where one dominant variant was replaced by the other one in our community. For instance, the Others (mainly B.1.2) were dominant until February 2021 and the Alpha variant raced for the dominant variant in around March 2021. Also, the Alpha variant was dominant from April and May in 2021, but the Delta variants competed with the Alpha variant in around June 2021 (**Fig. 4b**). Total SARS-CoV-2 concentrations (i.e., N gene concentrations) of the mixtures were 10^4^ and 10^1^ gc/µL, which are the reasonable concentration range of SARS-CoV-2 of local sewage samples (24). Also, we mixed the two different RNA controls with four different ratios (i.e., 9:1, 7:3, 3:7, and 1:9) to mimic different scenarios of variant dynamics.

### PCR assays for SARS-CoV-2 variant detection in sewage samples

We conducted six different RT-qPCR assays to analyze sewage samples. Three of them targeted different loci of SARS-CoV-2 genome for virus quantification and the dominant variant detection. The other three assays were applied to measure bovine coronavirus (BCoV), pepper mild mottle virus (PMMoV), and Tulane virus (TV), which were used for calculation of virus recovery efficiency, normalization of SARS-CoV-2 to human feces, and inhibition tests, respectively. 2-fold diluted the RNA samples in molecular biology grade water (Milliporesigma, USA) before the quantification. We used Taqman-based RT-qPCR for the N1 gene detection as suggested by CDC and SYBR-based RT-qPCR for the other six assays (**SI Table 6**). The SYBR-based RT-qPCR started with mixing the 3 μL of viral genome with 5 μL of 2 × iTaq universal SYBR green reaction mix, 0.125 μL of iScript reverse transcriptase from the iTaq universal SYBR green reaction mix (Bio-Rad Laboratories, USA), 0.3 μL of 10 μM forward primer for each virus, 0.3 μL of 10 μM reverse primer for each virus, and 1.275 μL of molecular biology grade water (Corning, NY, USA). The PCR cocktail for the one-step RT-qPCR was placed in 96-well plates (4306737, Applied Biosystems, USA) and analyzed by a qPCR system (QuantStudio 3, Thermo Fisher Scientific, USA). The thermocycle began with 10 minutes at 50°C and 1 minute at 90°C followed by 40 cycles of 30 seconds at 60°C and 1 minute at 90°C. The Taqman-based RT-qPCR was initiated by mixing 5 μL of viral genome with 5 μL of Taqman Fast Virus 1-step Master Mix (4444432, Applied Biosystems, USA), 1.5 μL of primers/probe mixture for N1 gene (2019-nCoV RUO kit, Integrated DNA Technologies, USA), and 8.5 μL of water. The 20 μL of mixture was analyzed by the same qPCR system as used for the SYBR-based RT-qPCR, except for a different thermal cycle (5 minutes at 50°C, 20 seconds at 95°C followed by 45 cycles of 3 seconds at 95°C and 30 seconds at 55°C). We used synthetic RNA controls to get standard curves for WT, Alpha variant, and Delta variant (TWIST Bioscience, USA, Part numbers are 102024, 103907, and 104533, respectively). The PCR standard curves were obtained for every RT-qPCR analysis with 10-fold serial dilutions of synthetic RNA controls and PCR efficiencies for RT-qPCR were higher than 85% (R^2^>0.99). The SYBR signal was normalized to the ROX reference dye. The cycle of quantification (Cq) values was determined automatically by QuantStudio™ Design & Analysis Software (v1.5.1). Based on the melting curves, the primers were specifically bound to the target genome. The numbers of technical replicates were 4 for synthetic RNA controls and 3 for sewage samples except for LODs and LOQs determination where 20 replicates were analyzed.

### Next-generation sequencing to assign SARS-CoV-2 lineages

The PCR results were confirmed by sequencing the spike gene of three controls (wild type, alpha variant, delta variant) and two sewage samples (#5 and #6) were performed on the Illumina MiSeq platform. A set of four pairs of primers in-house designed were used to amplify the spike of RNA samples using SuperScript(tm) III One-Step RT-PCR System with Platinum(tm) Taq High Fidelity DNA Polymerase (ThermoFisher). Amplicons were purified using QIAquick PCR Purification Kit (Qiagen), quantified using Qubit fluorometer, and subject to library preparation using Nextera XT kit and sequencing on MiSeq.

## Data Availability

All data produced in the present study are available upon reasonable request to the authors

## Data availability

All the sequence data analyzed in this study is publicly available at GISAID (https://www.gisaid.org/). The analyzed and processed real data results are available at https://github.com/elkebir-group/primes-data.

## Code availability

The code has been deposited on Github at https://github.com/elkebir-group/primes.

## Acknowledgement

We acknowledge the funding from the Grainger College of Engineering and the JUMP-ARCHES program of OSF Healthcare in conjunction with the University of Illinois. Sequencing was funded in part by the Food and Drug Administration Veterinary Laboratory Investigation and Response Network (FOA PAR-17-141) under grant 1U18FD006673-01. M.E-K. acknowledges the National Science Foundation (grants: CCF-2027669 and CCF-2046488). The authors also acknowledge Bill Brown for sampling site selection, Hayden Wennerdahl, Kip Stevenson, Dr. Laura Keefer and Dr. Schmidt for sampling deployment, and Yuqing Mao, Matthew Robert Loula, Aashna Patra, Kristin Joy Anderson, Mikayla Diedrick, Hubert Lyu, Hamza Elmahi Mohamed, Jad R Karajeh, Runsen Ning, Rui Fu, Kate O’Brien, Nathan Kim for sewage sampling and processing.

