## Supplemental Figures and Tables for "Regional and temporal variations affect the accuracy of variant-specific SARS-CoV-2 PCR assays"

**Six tables and six figures are included**

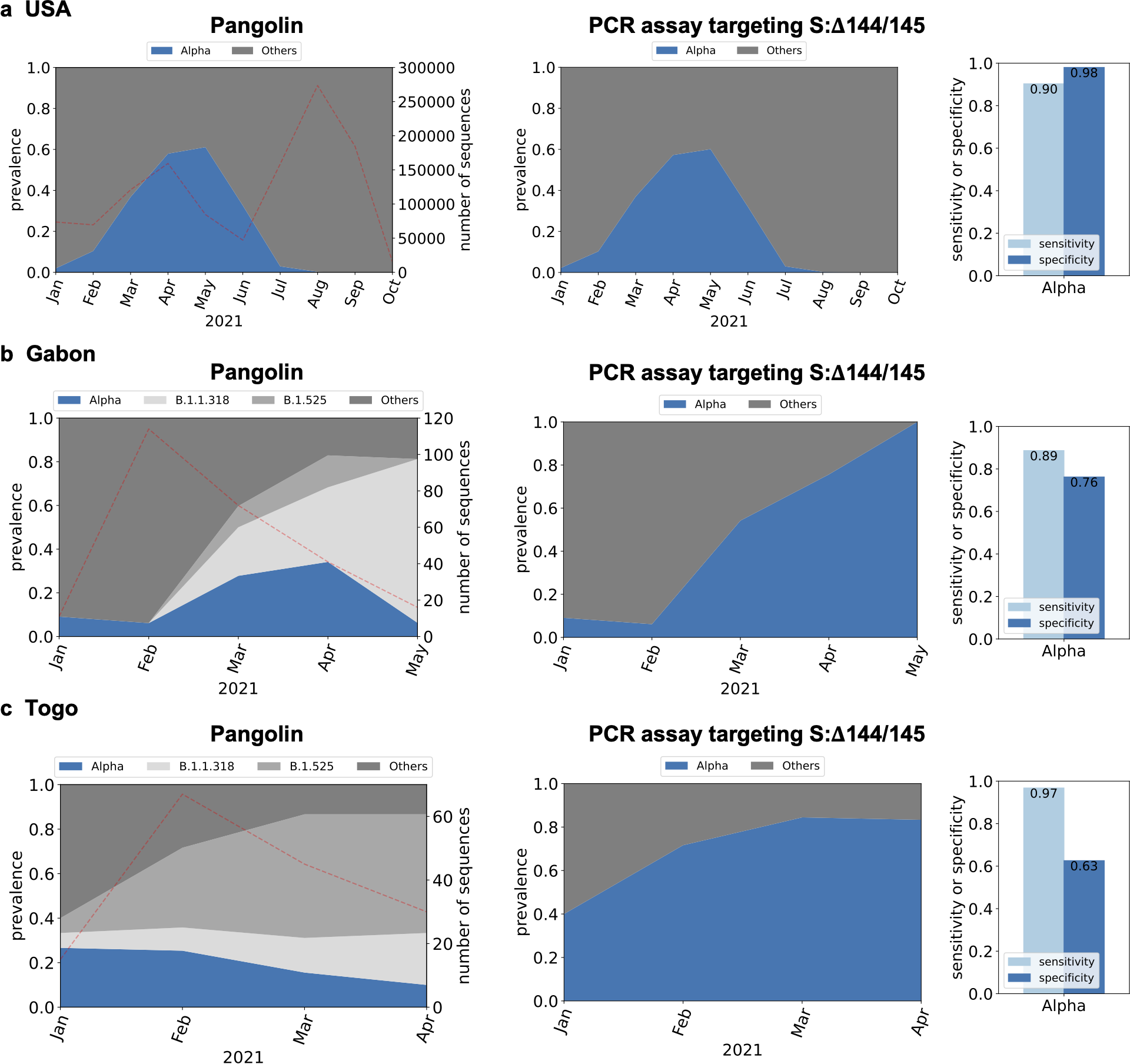

**SI Fig. 1.** *In silico* analysis of PCR assay targeting S:Δ144/145 mutation (1) to detect the Alpha variant for GISAID samples from (**a**) USA (n=1,187,412), (**b**) Gabon (n=254) and (**c**) Togo (n=157). Dotted lines on left figures indicate the number of sequences used for the *in silico* analyses.

**SI Table 1.** Defining mutations for Alpha, Gamma, and Delta variants with other lineages that cause false positives and false negatives (updated as of September 2021). If the predominance of target mutation in the target variant is less than 90%, lineages are considered to cause false positives in this study. If the predominance of target mutation in the other variants is higher than 10%, lineages are considered false negatives. Lineages with more than 100 sequences reported in GISAID are selectively listed.

| **Variant** | **Defining mutations** | **Analysis using Outbreak.info** |
| --- | --- | --- |
| Alpha | S:H69- | B.1.258.19, B.1.258.9, B.1.258.5, B.1.258.7, B.1.258.4, B.1.258.21, B.1.258.11 B.1.620, B.1.415.1, B.1.258.12, B.1.375, B.1.634, B.1.525 (Eta), A.28, B.1.1.298, B.1.258.22, C.36.3.1, C.36.3, B.1.625, B.1.258, B.1.1.189, B.1.1.205, B.1.636 (False positive)  Q.2 (False negative) |
|  | S:V70- |  |
|  | S:Y144- | AZ.6, B.1.620, AZ.2.1, AZ.2, AZ.4, B.1.525 (Eta), B.1.1.525, B.1.1.318, AZ.5, AZ.3, B.1.637, B.1.630, AV.1, C.1.2, B.1.625, B.1.160.24, AZ.1, B.1.177.80, B.1.1.189, B.1.636, C.38 (False positive) |
|  | S:N501Y | P.1.6, A29, P.1.4, A.27, B.1.621.1, P.1.3, P.1, AP.1, P.1.10, P.1.9, P.1.8, P.1.2, P.1.621, P.1.7, P.1.1, B.1.351.3, B.1.351.2, B.1.351, P.3, C.1.2, B.1.623, B.1.1.189, B.1.153, B.1.597, C.38, B.1.1.205, B.1.636 (False positive)  Q.5 (False negative) |
|  | **S:A570D** | B.1.1.189, C38, B.1.636 (False positive) |
|  | S:D614G | Too many lineages for false positive^1)^ |
|  | S:P681H | P.1.6, B.1.632, AZ.6, AZ.1, B.1.620, B.1.1.519, AZ.4, B.1.631, B.1.575.2, AZ.3, B.1.621.1, AZ.2, AZ.2.1, B.1.1.351, B.1.1.318, B.1.1.526, B.1.621, B.1.415.1, P.3, AZ.5, B.1.628, B.1.575.1, B.1.627, B.1.1.207, B.1.474, P.1.7, AV.1, B.1.469, B.1.575, B.1.243, B.1.468, B.1.400, P.1.1., B.1.189, B.1.1.274, B.1.1.485, B.1.636, B.1.1.163, A.19, B.1.1.161, C.38, B.1.1.46, B.1.1.263 (False positive)  Q.4, Q.5, Q.8 (False negative) |
|  | S:T716I | B.1.214.4, B.1.575.2, B.1.214.2, B.1.575.1, C.1.2, B.1.214.3, B.1.575, B.1.1.205, B.1.214, B.1.1.377 (False positive)  Q5, Q8 (False negative) |
|  | S:S982A | Q8 (False negative) |
|  | S:D1118H | B.1.620, B.1.1.189 (False positive) |
|  | ORF1a:T1001I | B.1.36.29, AB.1, B.1.636, B.1.1.189, B.1.456 (False positive) |
|  | ORF1a:A1708D | B.1.1.189, B.1.1.205, B.1.636 (False positive) |
|  | ORF1a:I2230T | B.1.1.189, B.1.1.205, B.1.1.226, B.1.636 (False positive) |
|  | ORF1a:S3675- | P.1.6, AZ.6, P.1.10.1, P.1.10.2, P.1.5, AZ.1, B.1.620, B.1.3, B.1.630, P.1.8, B.1.351.2, P.1.9, B.1.619.1, P.1.4, AZ.2, B.1.526, AZ.2.1, P.1.2, B.1.619, B.1.636, P.1.10, B.1.637, AV.1, B.1.1.318, B.1.634, P.1, P.1.7, C.37.1, B.1.1.525, C.37, B.1.351.3, B.1.525, AZ.3, C.1.2, B.1.1.528, AZ.4, B.1.625, B.1.351, B.1.628, P.1.11, P.1.1, AZ.5, B.1.214.2, B.1.470, B.1.1.189, C.38, B.1.1.205, B.1.241, B.1.214.3, B.1.237, B.1.153, B.1.1.375 (False positive) |
|  | ORF1a:G3676- |  |
|  | ORF1a:F3677- |  |
|  | N:D3L | B.1.214.4, B.1.214.2, B.1.214.3, B.1.1.189, B.1.214 (False positive) |
|  | N:R203K | Too many lineages for false positive^2)^  Q.5 (False negative) |
|  | N:G204R | Too many lineages for false positive^3)^  Q.5, Q.6, Q.7 (False negative) |
|  | N:S235F | B.1.565, B.1.139, B.1.170, C.38, B.1.1.226, B.1.533, B.1.378 (False positive) |
|  | ORF1b:P314L | Too many lineages for false positive^4)^  Q.8 (False negative) |
|  | ORF8:Q27* | B.1.1.189, B.1.221.4, B.1.636 (False positive)  Q.5, Q.2 (False negative) |
|  | ORF8:R52I | B.1.636, B.1.1.189 (False positive)  Q.5, Q.2 (False negative) |
|  | ORF8:Y73C | B.1.1.189, B.1.1.205 (False positive) |
| Gamma | **S:L18F** | No lineages are discovered for false positives and false negatives |
|  | **S:T20N** |  |
|  | S:P26S | B.1.620, B.1.36.36, B.1.1.420, B.1.177.57 (False positive) |
|  | S:D138Y | B.1.1.333, B.1.561, B.1.1.413, B.1.1.397, B.1.1.354, B.1.1.528, B.1.1.317, B.1.428, B.1.241, B.1.560, B.1.36.24 (False positive)  P.1.3 (False negative) |
|  | S:R190S | C.1.2, A.21 (False positive)  P.1.2 (False negative) |
|  | S:K417T | P.1.10.2 (False negative) |
|  | S:E484K | AZ.6, B.1.619.1, AZ.2, R.1, B.1.620, AZ.1, B.1.575.2, B.1.626, N.9, AZ.2.1, AZ.3, B.1.1.318, B.1.525, B.1.632, B.1.621.1, B.1.618, AZ.4, B.1.634, B.1.1.523, B.1.619, B.1.621, AZ.5, B.1.1.525, B.1.351.3, AT.1, B.1.351.2, B.1.625, B.1.351, C.1.2, AV.1, B.1.415.1, C.38, B.1.526, B.1.1.207, B.1.1.526 (False positive) |
|  | S:N501Y | A.29, Q.1, Q.7, Q.2, Q.6, Q.3, A.27, B.1.1.7, B.1.621.1, Q.4, AP.1, B.1.621, Q.8, B.1.351.3, B.1.351.2, B.1.351, Q.5, P.3, C.1.2, B.1.623, B.1.1.189, B.1.153, B.1.597, C.38, B.1.1.205, B.1.636 (False positive) |
|  | S:D614G | Too many lineages for false positive^1)^ |
|  | S:H655Y | B.1.160.7, B.1.160.33, A.29, B.1.632, A.28, A.27, B.1.630, B.1.1.525, C.1.2, B.1.1.157, B.1.438 (False positive) |
|  | S:T1027I | B.1.619.1, B.1.631, B.1.620, B.1.619, B.1.1.523, B.1.628, B.1.627, B.1.636 (False positive)  P.1.1 (False negative) |
|  | S:V1176F | P.4, P.6, P.7, P.2, B.1.1.28, P.3, B.1.324, B.1.177.75 (False positive)  P.1.1 (False negative) |
|  | ORF3a:S253P | No lineages are discovered for false positives and false negatives |
|  | ORF1a:S1188L | No lineages are discovered for false positives and false negatives |
|  | ORF1a:K1795Q | B.1.630 (False positive) |
|  | ORF1a:S3675- | AZ.6, AZ.1, B.1.620, Q.7, B.1.630, Q.8, B.1.351.2, B.1.619.1, AZ.2, B.1.526, AZ.2.1, B.1.619, Q.1, Q.4, B.1.636, B.1.637, Q.6, AV.1, B.1.1.318, B.1.634, Q.3, B.1.1.7, C.37.1, B.1.1.525, C.37, B.1.351.3, B.1.525, AZ.3, C.1.2, B.1.1.528, AZ.4, B.1.625, B.1.351, B.1.628, Q.5, AZ.5, B.1.214.2, B.1.470, Q.2, B.1.1.189, C.38, B.1.1.205, B.1.241, B.1.214.3, B.1.237, B.1.153, B.1.1.375 (False positive) |
|  | ORF1a:G3676- |  |
|  | ORF1a:F3677- |  |
|  | N:P80R | B.1.630 (False positive) |
|  | N:R203K | Too many lineages for false positive^5)^ |
|  | N:G204R | Too many lineages for false positive^6)^ |
|  | ORF1b:P314L | Too many lineages for false positive^7)^ |
|  | ORF1b:E1264D | B.1.1.272, B.1.375, B.1.36.38 (False positive) |
|  | ORF8:E92K | A.23.1, A.29, A (False positive) |
| Delta | S:T19R | B.1.617.3, B.1.1.374, B.1.623 (False positive)  AY.28 (False negative) |
|  | S:E156_ | AY.13, AY.5.2, AY.14, AY.21, AY.38, AY.35, AY.15, AY.10, AY.23, AY.19, AY.36, AY.26, AY.16, AY.32, AY.30, AY.1, AY.24, AY.37, AY.17, AY.7.1, AY.33 (False negative) |
|  | S:F157_ |  |
|  | S:L452R | P.4, C.36.3.1, A.2.5.1, B.1.637, B.1.429, A.27, B.1.427, C.36.3, A.2.5.2, C.16, B.1.617.1, B.1.630, L.3, A.2.5, B.1.1.487, B.1.617.3, B.1.36.35, A.21, B.1.362, B.1.623, C.36, B.1.177.83, B.1.232, B.1.1.354, B.1.459, B.1.575 (False positive)  AY.32, AY.28, AY.10 (False negative) |
|  | S:T478K | B.1.1.519, B.1.214.3, B.1.36.1, B.1.623, C.1.2 (False positive)  AY.32, AY.28, AY.10 (False negative) |
|  | S:D614G | Too many lineages for false positive^1)^ |
|  | **S:P681R** | AU.3, AU.2, P.1.8, B.1.617.3, A.23.1, B.1.617.1, B.1.551, B.1.466.2, B.1.1.528, Q.4, B.1.623, B.1.1.25, C.36 (False positive)  AY.28 (False negative) |
|  | S:D950N | B.1.621.1, B.1.630, B.1.621, B.1.625, B.1.617.3, B.1.623 (False positive)  AY.20, AY.26, AY.16, AY.19, AY.7.2, AY.32, AY.28 (False negative) |
|  | ORF1b:P314L | Too many lineages for false positive^7)^ |
|  | ORF1b:P1000L | A.2.5.1, B.1.634, A.2.5.2, A.27, B.1.1.523, A.25, B.1.628, B.1.623, B.1.459 (False positive)  AY.16, AY.28 (False negative) |
|  | M:I82T | AZ.1, AZ.6, B.1.619.1, AZ.4, C.36.3.1, AZ.2, B.1.632, AZ.3, AZ.2.1, B.1.575.2, B.1.525, B.1.1.318, C.36.3, AZ.5, B.1.575.1, B.1.1.523, B.1.415.1, B.1.619, B.1.625, B.1.575, C.1.2, B.1.1.528, C.38, B.1.623 (False positive)  AY.32 (False negative) |
|  | N:D63G | B.1.623 (False positive)  AY.7.2, AY.23, AY.18 (False negative) |
|  | N:R203M | B.1.617.1, B.1.617.3, B.1.623, B.1.189, B.1.459 (False positive)  AY.24, AY.23 (False negative) |
|  | N:D377Y | B.1.1.288, B.1.1.368, B.1.367, B.1617.1, B.1.617.3, B.1.433, B.1.1.229, B.1.36.19, B.1.177.42, B.1.110.3, B.1.177.46, S.1, B.1.258.3, B.1.459, B.1.2, A.2.5.1, B.1.22, B.1.214.3, B.1.623, L.3, B.1.631, AD.2 (False positive)  AY.12 (False negative) |
|  | ORF3a:S26L | B.1.617.1, B.1.623 (False positive) |
|  | ORF7a:V82A | B.1.617.3, B.1.617.1, B.1.623, B.1.459 (False positive)  AY.7.1, AY.24, AY.23, AY.7.2, AY.27, AY.32, AY.16, AY.17, AY.12 (False negative) |
|  | ORF7a:T120I | B.1.177.18, Q.2, B.1.160.31, B.1.1.58, B.1.623 (False positive)  AY.7.1, AY.23, AY.27, AY.16, AY.12 (False negative) |

1) <https://outbreak.info/situation-reports?pango&muts=S%3AD614G> (Last access on September 30th, 2021)
2) <https://outbreak.info/situation-reports?pango&muts=N%3AR203K> (Last access on September 30th, 2021)
3) <https://outbreak.info/situation-reports?pango&muts=N%3AG204R> (Last access on September 30th, 2021)
4) <https://outbreak.info/situation-reports?pango&muts=ORF1b%3AP314L> (Last access on September 30th, 2021)
5) <https://outbreak.info/situation-reports?pango&muts=N%3AR203K> (Last access on September 30th, 2021)
6) <https://outbreak.info/situation-reports?pango&muts=N%3AG204R> (Last access on September 30th, 2021)
7) <https://outbreak.info/situation-reports?pango&muts=ORF1b%3AP314L> (Last access on September 30th, 2021)
8) The up-to-date data will be found via (https://outbreak.info/)

**Table 2.** Predominance of various sub-lineages in different countries. The countries with higher than 1% predominance for each sub-lineage are reported. The predominance was determined by Outbreak.info. The predominance was determined in Sep. 2021, and the latest predominance should be checked via Outbreak.info.

| Sub-lineage | Country | Prevalence (target sequences/total sequences) |
| --- | --- | --- |
| A | Too many countries | - |
| A.19 | Côte d'Ivoire | 20% (48/236) |
|  | Burkina Faso | 12% (48/403) |
| A.21 | Burkina Faso | 21% (84/404) |
|  | Mali | 4% (3/71) |
|  | Côte d'Ivoire | 3% (7/236) |
|  | Central African Republic | 2% (1/56) |
| A.23.1 | Rwanda | 27% (129/483) |
|  | Uganda | 25% (160/628) |
|  | South Sudan | 10% (9/88) |
|  | Central African Republic | 2% (1/56) |
|  | Democratic Republic of the Congo | 2% (11/649) |
|  | Benin | 2% (1/65) |
|  | Kenya | 1% (48/3341) |
|  | Vietnam | 1% (5/357) |
|  | Cambodia | 1% (10/992) |
| A.27 | Côte d'Ivoire | 10% (24/236) |
|  | Togo | 7% (23/343) |
|  | Tunisia | 5% (7/129) |
|  | Burkina Faso | 5% (21/403) |
|  | Benin | 3% (9/263) |
|  | Algeria | 2% (1/54) |
|  | Mayotte | 2% (12/727) |
| A.28 | Canary Islands | 9% (34/358) |
|  | Egypt | 2% (21/993) |
| A.29 | Sudan | 18% (17/97) |
| AT.1 | Crimea | 3% (1/37) |
|  | Russia | 1% (113/8195) |
| AV.1 | Namibia | 1% (3/262) |
|  | Belarus | 1% (1/90) |
| AY.1 | Nepal | 5% (12/257) |
|  | Albania | 2% (1/42) |
|  | Georgia | 1% (3/258) |
| AY.5.2 | Portugal | 1% (195/18011) |
| AY.7.1 | Anguilla | 27% (3/11) |
|  | Antigua and Barbuda | 23% (14/)60 |
|  | Armenia | 14% (20/140) |
|  | Saint Vincent and the Grenadines | 7% (1/14) |
|  | Seychelles | 7% (18/256) |
|  | Barbados | 6% (5/77) |
|  | Turks and Caicos Islands | 6% (1/16) |
|  | Swaziland | 6% (7/121) |
|  | Ghana | 6% (81/1422) |
|  | Liechtenstein | 6% (5/90) |
|  | Denmark | 4% (6942/171912) |
|  | Jamaica | 3% (5/177) |
|  | Kuwait | 2% (7/283) |
|  | Equatorial Guinea | 2% (5/205) |
|  | Kosovo | 2% (10/483/) |
|  | Lebanon | 2% (16/1019) |
|  | Morocco | 1% (5/387) |
| AY.10 | Gambia | 8% (51/608) |
|  | Senegal | 1% (6/513) |
|  | Uzbekistan | 1% (1/88) |
| AY.11 | Anguila | 9% (1/11) |
|  | Barbados | 3% (2/78) |
| AY.13 | U.S. Virgin Islands | 5% (18/365) |
| AY.14 | Turks and Caicos Islands | 6% (1/16) |
| AY.16 | Kenya | 11% (379/3513) |
|  | Kazakhstan | 9% (43/460) |
|  | Crimea | 8% (3/37) |
|  | Afghanistan | 5% (5/94) |
|  | Andorra | 4% (1/25) |
|  | South Sudan | 3% (3/88) |
|  | Nepal | 2% (4/257) |
|  | Mali | 1% (1/71) |
|  | Republic of Congo | 1% (9/651) |
|  | Oman | 1% (11/864) |
|  | India | 1% (726/61325) |
|  | Uzbekistan | 1% (1/88) |
|  | Gibraltar | 1% (17/1648) |
| AY.17 | Nepal | 2% (4/257) |
| AY.19 | Swaziland | 2% (2/121) |
|  | Kuwait | 1% (3/283) |
| AY.21 | Nepal | 2% (4/257) |
|  | Lebanon | 1% (11/1019) |
| AY.23 | Singapore | 54% (4283/7919) |
|  | Brunei | 37% (13/35) |
|  | Indonesia | 32% (2238/7099) |
|  | Malaysia | 14% (454/3252) |
|  | Timor-Leste | 9% (31/353) |
|  | Malta | 6% (15/256) |
|  | Barbados | 4% (3/77) |
|  | Paraguay | 3% (10/386) |
|  | Papua New Guinea | 2% (8/332) |
|  | Albania | 2% (1/42) |
|  | Seychelles | 2% (5/256) |
|  | Nepal | 2% (4/257) |
|  | Mauritius | 2% (4/266) |
|  | Kosovo | 1% (7/483) |
|  | Ethiopia | 1% (2/140) |
|  | Kazakhstan | 1% (5/460) |
| AY.24 | Brunei | 43% (15/35) |
|  | Indonesia | 10% (710/7099) |
|  | Kosovo | 1% (5/483) |
| AY.26 | Bahrain | 28% (247/873) |
|  | Mexico | 10% (3180/30913) |
|  | Uzbekistan | 10% (9/88) |
|  | Anguilla | 9% (1/11) |
|  | Bangladesh | 3% (91/2791) |
|  | India | 3% (1771/61325) |
|  | Albania | 2% (1/42) |
|  | Czech Republic | 2% (204/9129) |
|  | U.S. Virgin Islands | 2% (7/365) |
|  | Costa Rica | 2% (24/1273) |
|  | United States | 2% (20332/1180345) |
|  | Swaziland | 2% (2/121) |
|  | Puerto Rico | 2% (52/3232) |
|  | Kuwait | 1% (4/283) |
|  | Fiji | 1% (7/531) |
|  | Liberia | 1% (1/77) |
|  | Malta | 1% (3/256) |
|  | Sri Lanka | 1% (16/1370) |
|  | Iceland | 1% (109/9590) |
|  | Malaysia | 1% (33/3252) |
| AY.30 | Australia | 29% (9955/34066) |
|  | Thailand | 19% (682/3638) |
|  | Cambodia | 18% (212/1163) |
|  | Myanmar | 9% (7/75) |
|  | Grenada | 8% (1/12) |
|  | Kosovo | 1% (5/483) |
| AY.32 | South Africa | 4% (895/19980) |
|  | Mauritius | 3% (9/266) |
|  | Ethiopia | 1% (2/140) |
|  | Bahrain | 1% (9/873) |
| AY.33 | Maldives | 67% (173/258) |
|  | Botswana | 44% (461/1036) |
|  | Nepal | 27% (70/257) |
|  | Morocco | 10% (39/387) |
|  | India | 8% (5160/61325) |
|  | Armenia | 5% (7/140) |
|  | Bahrain | 3% (22/873) |
|  | Ethiopia | 2% (3/140) |
|  | Namibia | 2% (6/282) |
|  | Costa Rica | 2% (27/1273) |
|  | Togo | 2% (7/343) |
|  | Switzerland | 2% (1370/68952) |
|  | Seychelles | 2% (5/256) |
|  | Belgium | 2% (932/50135) |
|  | Kenya | 2% (63/3513) |
|  | Swaziland | 2% (2/121) |
|  | Kuwait | 1% (4/283) |
|  | Germany | 1% (2750/206790) |
|  | Monaco | 1% (1/78) |
|  | Zambia | 1% (11/887) |
|  | Uzbekistan | 1% (1/88) |
|  | Denmark | 1% (1743/171912) |
| AY.35 | British Virgin Islands | 3% (1/33) |
|  | Jamaica | 1% (2/177) |
|  | Puerto Rico | 1% (35/3232) |
| AY.36 | Nigeria | 35% (936/2701) |
|  | Burkina Faso | 5% (21/404) |
|  | Maldives | 4% (10/258) |
|  | Benin | 2% (4/262) |
| AY.37 | Liberia | 65% (50/77) |
|  | Canada | 3% (3990156132/) |
|  | Kuwait | 2% (6/283) |
|  | Monaco | 1% (1/78) |
|  | Seychelles | 1% (3/256) |
|  | U.S. Virgin Islands | 1% (4/365) |
| AY.38 | Swaziland | 6% (7/121) |
|  | South Africa | 5% (941/19980) |
|  | Georgia | 2% (4/258) |
| AZ.1 | Cayman Islands | 1% (1/73) |
| AZ.2 | Greece | 9% (867/9336) |
|  | Togo | 1% (5/343) |
| AZ.2.1 | Liechtenstein | 12% (13/109) |
| AZ.4 | Ireland | 1% (284/32538) |
| AZ.5 | Mauritius | 72% (192/266) |
| B.1.1.7 | Too many countries | - |
| B.1.1.28 | Paraguay | 11% (42/386) |
|  | Philippines | 6% (398/7036) |
|  | brazil | 4% (1934/46237) |
|  | uruguay | 3% (18/719) |
| B.1.177.57 | Solomon Islands | 67% (4/6) |
| B.1.177.80 | Iraq | 1% (2/169) |
| B.1.1.189 | Bolivia | 2% (1/66) |
| B.1.1.298 | Faroe Islands | 2% (1/42) |
| B.1.1.317 | Russia | 7% (605/8195) |
|  | Faroe Islands | 2% (1/42) |
|  | Estonia | 2% (119/6218) |
|  | China | 1% (14/1008) |
|  | Belarus | 1% (1/87) |
|  | Uzbekistan | 1% (1/88) |
| B.1.1.318 | Benin | 17% (11/65) |
|  | Gabon | 16% (42/267) |
|  | Ghana | 13% (176/1337) |
|  | Togo | 10% (34/343) |
|  | Nigeria | 5% (95/1815) |
|  | Liberia | 4% (3/77) |
|  | Guinea | 4% (8/214) |
|  | Antigua and Barbuda | 2% (1/60) |
|  | Bangladesh | 1% (32/2548) |
|  | Cameroon | 1% (2/197) |
| B.1.1.333 | Norway | 3% (874.27634) |
| B.1.1.354 | Brunei | 6% (2/35) |
| B.1.1.375 | Mozambique | 11% (63/574) |
| B.1.1.397 | Mongolia | 4% (1/28) |
|  | Russia | 3% (206/8195) |
| B.1.1.413 | Azerbaijan | 7% (1/14) |
|  | Iran | 5% (30/552) |
| B.1.1.420 | Cabo Verde | 15% (6/39) |
|  | Guinea-Bissau | 15% (7/48) |
|  | Senegal | 14% (71/513) |
|  | Gambia | 2% (15/608) |
|  | Luxembourg | 1% (133/12737) |
| B.1.1.519 | Mexico | 29% (7752/27035) |
|  | Belize | 4% (2/52) |
|  | Martinique | 3% (12/387) |
|  | United States | 1% (13966/984576) |
|  | Kosovo | 1% (1/81) |
|  | Aruba | 1% (27/2524) |
|  | Guatemala | 1% (7/681) |
|  | Curaçao | 1% (7/681) |
| B.1.1.523 | Russia | 5% (383/8195) |
|  | Guam | 4% (8/196) |
|  | Moldova | 1% (1/67) |
| B.1.1.525 | Russia | 1% (96/8041) |
| B.1.36.36 | Canada | 1% (1639/156132) |
| B.1.170 | Sudan | 2% (2/97) |
| B.1.214.2 | Republic of the Congo | 22% (48/221) |
|  | Liechtenstein | 11% (11/107) |
|  | Democratic Republic of the Congo | 2% (13/649) |
|  | Angola | 2% (14/860) |
|  | Belgium | 2% (737/45463) |
| B.1.237 | Lesotho | 6% (1/18) |
| B.1.241 | Costa Rica | 1% (17/1273) |
| B.1.258 | Cyprus | 54% (72/134) |
|  | Liechtenstein | 25% (27/109) |
|  | Faroe Islands | 24% (10/42) |
|  | Czech Republic | 7% (577/8537) |
|  | Croatia | 4% (318/7417) |
|  | Montenegro | 4% (8/193) |
|  | Romania | 4% (92/2246) |
|  | Bosnia and Herzegovina | 3% (7/207) |
|  | Estonia | 3% (164/5861) |
|  | Finland | 2% (420/18157) |
|  | Slovenia | 2% (435/22059) |
|  | Canary Islands | 2% (7/358) |
|  | Iceland | 2% (183/9603) |
|  | Slovakia | 2% (118/6991) |
|  | Macedonia | 2% (11/699) |
|  | Poland | 2% (318/20532) |
|  | Hungary | 1% (6/435) |
|  | Germany | 1% (2645/196466) |
|  | Denmark | 1% (1998/168597) |
|  | Switzerland | 1% (742/66019) |
|  | Sweden | 1% (1127/105392) |
|  | Norway | 1% (275/26268) |
| B.1.258.11 | Denmark | 1% (2468/168597) |
| B.1.258.17 | Slovenia | 24% (5313/22059) |
|  | Macedonia | 5% (37/699) |
|  | Croatia | 2% 167/7417 |
|  | Liechtenstein | 2% (2/109) |
|  | Bosnia and Herzegovina | 1% (3/207) |
| B.1.324 | Djibouti | 4% (12/305) |
|  | Northern Mariana Islands | 2% (3/152) |
| B.1.378 | The Bahamas | 2% (2/133) |
| B.1.428 | Qatar | 22% (616/2764) |
|  | Myanmar | 1% (1/75) |
| B.1.438 | Iraq | 1% (3/220) |
| B.1.470 | Indonesia | 7% (485/7099) |
|  | Malaysia | 1% (41/3252) |
| B.1.525 | Libya | 50% (11/22) |
|  | South Sudan | 50% (44/88) |
|  | Mali | 44% (31/71) |
|  | Niger | 25% (6/24) |
|  | Togo | 20% (68/343) |
|  | Côte d'Ivoire | 19% (45/236) |
|  | Benin | 18% (12/65) |
|  | Nigeria | 16% (390/2509) |
|  | Liberia | 8% (6/77) |
|  | Burkina Faso | 6% (24/403) |
|  | Uganda | 6% (37/626) |
|  | Cameroon | 6% (11/200) |
|  | Ghana | 5% (66/1341) |
|  | Gabon | 5% (13/267) |
|  | Malta | 4% (11/253) |
|  | Guinea | 3% (6/227) |
|  | Kuwait | 2% (6/262) |
|  | Senegal | 2% (9/491) |
|  | Central African Republic | 2% (1/56) |
|  | Moldova | 1% (1/67) |
|  | Canada | 1% (1828/141562) |
|  | Angola | 1% (11/866) |
|  | Belarus | 1% (1/90) |
| B.1.533 | Saudi Arabia | 1% (11/1093) |
| B.1.561 | Guam | 1% (2/196) |
| B.1.597 | Algeria | 7% (4/54) |
|  | Tunisia | 2% (3/128) |
|  | Morocco | 2% (8/387) |
| B.1.621.1 | British Virgin Islands | 64% (21/33) |
|  | Dominican Republic | 19% (62/325) |
|  | Colombia | 6% (226/3839) |
|  | Haiti | 5% (5/95) |
|  | Ecuador | 2% (51/2202) |
|  | U.S Virgin Islands | 1% (5/365) |
|  | Barbados | 1% (1/78) |
| B.1.617.3 | Malawi | 2% (11/502) |
| B.1.620 | Central African Republic | 38% (21/56) |
|  | Republic of the Congo | 24% (54/226) |
|  | South Korea | 3% (390/13629) |
|  | Cameroon | 3% (6/200) |
|  | Gabon | 1% (3/267) |
| B.1.625 | Colombia | 6% (203/3544) |
|  | Dominican Republic | 2% (5/325) |
|  | Venezuela | 1% (2/170) |
| B.1.630 | Dominican Republic | 8% (26/325) |
|  | Turks and Caicos Islands | 6% (1/16) |
| B.1.634 | Honduras | 2% (2/116) |
|  | Cayman Islands | 1% (1/73) |
| B.1.636 | Honduras | 6% (7/116) |
| B.1.637 | Grenada | 8% (1/12) |
|  | Dominican Republic | 7% (22/325) |
|  | Puerto Rico | 4% (133/2993) |
|  | Suriname | 3% (16/627) |
|  | Cayman Islands | 1% (1/73) |
|  | Jamaica | 1% (2/177) |
|  | United States | 1% (12319/1114605) |
| C.1.2 | Swaziland | 5% (6/121) |
| C.36.3 | Egypt | 10% (97/993) |
|  | Belarus | 2% (1/51) |
|  | Saudi Arabia | 2% (17/1096) |
| C.38 | Egypt | 1% (11/993) |
| P.1 | Trinidad and Tobago | 62% (357/579) |
|  | Suriname | 55% (342/627) |
|  | Brazil | 52% (23130/44649) |
|  | Haiti | 48% (46/95) |
|  | French Guiana | 48% (396/819) |
|  | Chile | 38% (3976/10404) |
|  | Guyana | 29% (4/14) |
|  | Bolivia | 26% (17/66) |
|  | Montserrat | 25% (1/4) |
|  | Uruguay | 23% (169/720) |
|  | Paraguay | 23% (87/386) |
|  | Colombia | 16% (597/3839) |
|  | Argentina | 15% (1093/7515) |
|  | Dominican Republic | 12% (38/325) |
|  | Ecuador | 11% (245/2202) |
|  | Costa Rica | 11% (135/1273) |
|  | Canada | 10% (14706/145942) |
|  | Venezuela | 10% (17/170) |
|  | Malta | 9% (24/253) |
|  | Mexico | 8% (2431/29445) |
|  | Luxembourg | 8% (1000/12752) |
|  | Peru | 7% (453/6499) |
|  | Barbados | 6% (5/78) |
|  | Antigua and Barbuda | 5% (3/60) |
|  | Belize | 4% (7/178) |
|  | Belgium | 4% (1769/48897) |
|  | Guatemala | 4% (24/683) |
|  | Faroe Islands | 2% (1/42) |
|  | United States | 2% (22726/1144329( |
|  | Curaçao | 2% (14/707) |
|  | Taiwan | 2% (4/239) |
|  | Aruba | 2% (39/2442) |
|  | Spain | 2% (1031/68050) |
|  | Puerto Rico | 1% (47/3213) |
|  | Montenegro | 1% (3/216) |
|  | Cayman Islands | 1% (1/73) |
|  | Italy | 1% (802/60829) |
|  | Jordan | 1% (10/779) |
|  | Portugal | 1% (189/17493) |
| P.1.1 | Malta | 3% (8/253) |
|  | Italy | 3% (1716/60829) |
|  | Suriname | 2% (13/627) |
|  | Trinidad and Tobago | 1% (6/579) |
| P.1.2 | Argentina | 2% (143/7515) |
|  | Brazil | 2% (693/44649) |
|  | Haiti | 1% (1/95) |
| P.1.4 | Brazil | 1% (590/44649) |
| P.1.7 | Peru | 7% (454/6499) |
|  | Brazil | 4% (1667/44649) |
|  | Haiti | 2% (2/95) |
|  | Chile | 1% (60/10404) |
| P.1.10 | Haiti | 7% (7/95) |
|  | Honduras | 2% (2/116) |
| P.2 | Suriname | 15% (93/626) |
|  | paraguay | 10% (38/386) |
|  | french Guiana | 9% (70/819) |
|  | Uruguay | 6% (41/719) |
|  | brazil | 6% (2601/46237) |
|  | canary Islands | 6% (24/432) |
|  | argentina | 1% (90/7567) |
| P.3 | Philippines | 4% (212/5345) |
|  | Guam | 2% (3/196) |
| P.6 | Uruguay | 39% (283/719) |
|  | Azerbaijan | 7% (1/14) |
| P.7 | Uruguay | 4% (28/719) |
|  | Paraguay | 3% (10/386) |
| Q.1 | Lithuania | 25% (4884/19796) |
|  | Belarus | 2% (2/87) |
|  | Japan | 1% (1650/129656) |
|  | Georgia | 1% (3/258) |
| Q.2 | Italy | 3% (1796/58744) |
| Q.4 | Georgia | 1% (3/256) |
|  | Sudan | 1% (1/97) |
| Q.7 | Oman | 1% (9/864) |
| Q.8 | Sri Lanka | 21% (286/1368) |
| R.1 | Sierra Leone | 37% (18/49) |
|  | Japan | 6% (7200/129656) |
|  | Guinea | 3% (7/223) |
|  | British Virgin Islands | 3% (1/33) |
|  | Trindad and Tobago | 3% (16/584) |
|  | Liberia | 1% (1/77) |
|  | Unknown | 1% (18/2845) |
|  | Ecuador | 11% (234/2202) |
|  | Haiti | 6% (6/95) |
|  | Argentina | 6% (455/7567) |
|  | Cameroon | 3% (7/202) |
|  | El Salvador | 3% (3/104) |
|  | Colombia | 2% (72/4116) |
|  | Bolivia | 2% (1/66) |
|  | Venezuela | 1% (2/171) |
|  | Costa Rica | 1% (14/1273) |

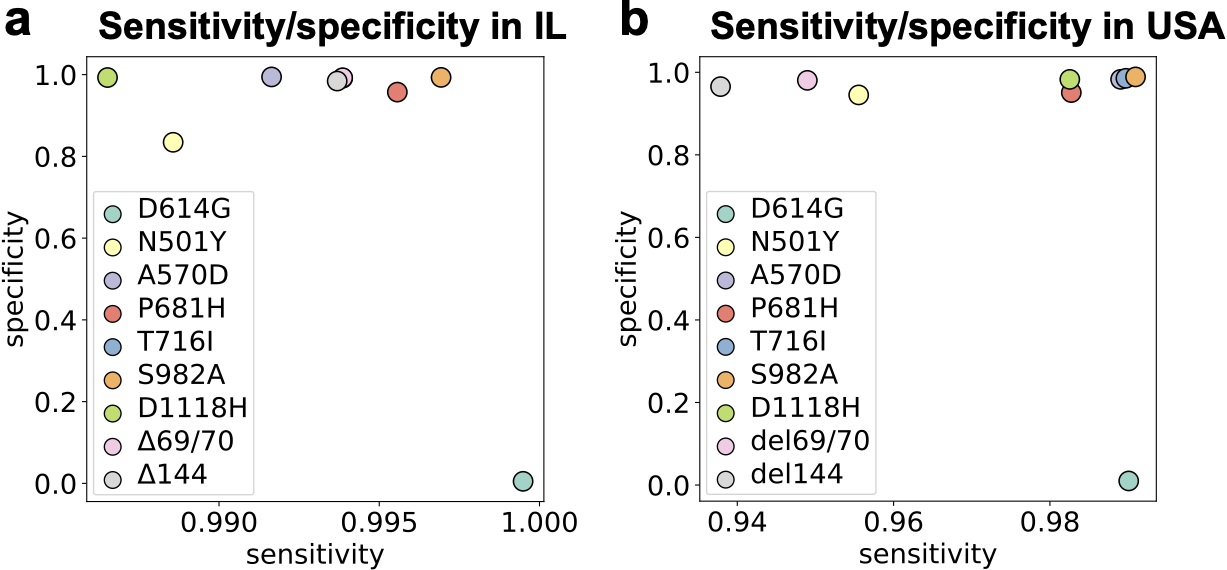
**SI Fig. 2.** Sensitivity and specificity of assigning the Alpha variant based on the presence of spike gene mutations characterizing the Alpha variant in GISAID samples from **(a)** IL and **(b)** USA.

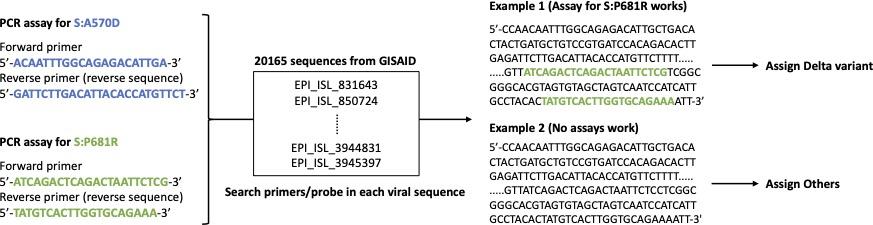

**SI Fig. 3.** An illustrative example showing the assignment of lineages using PCR assays designed in this study.

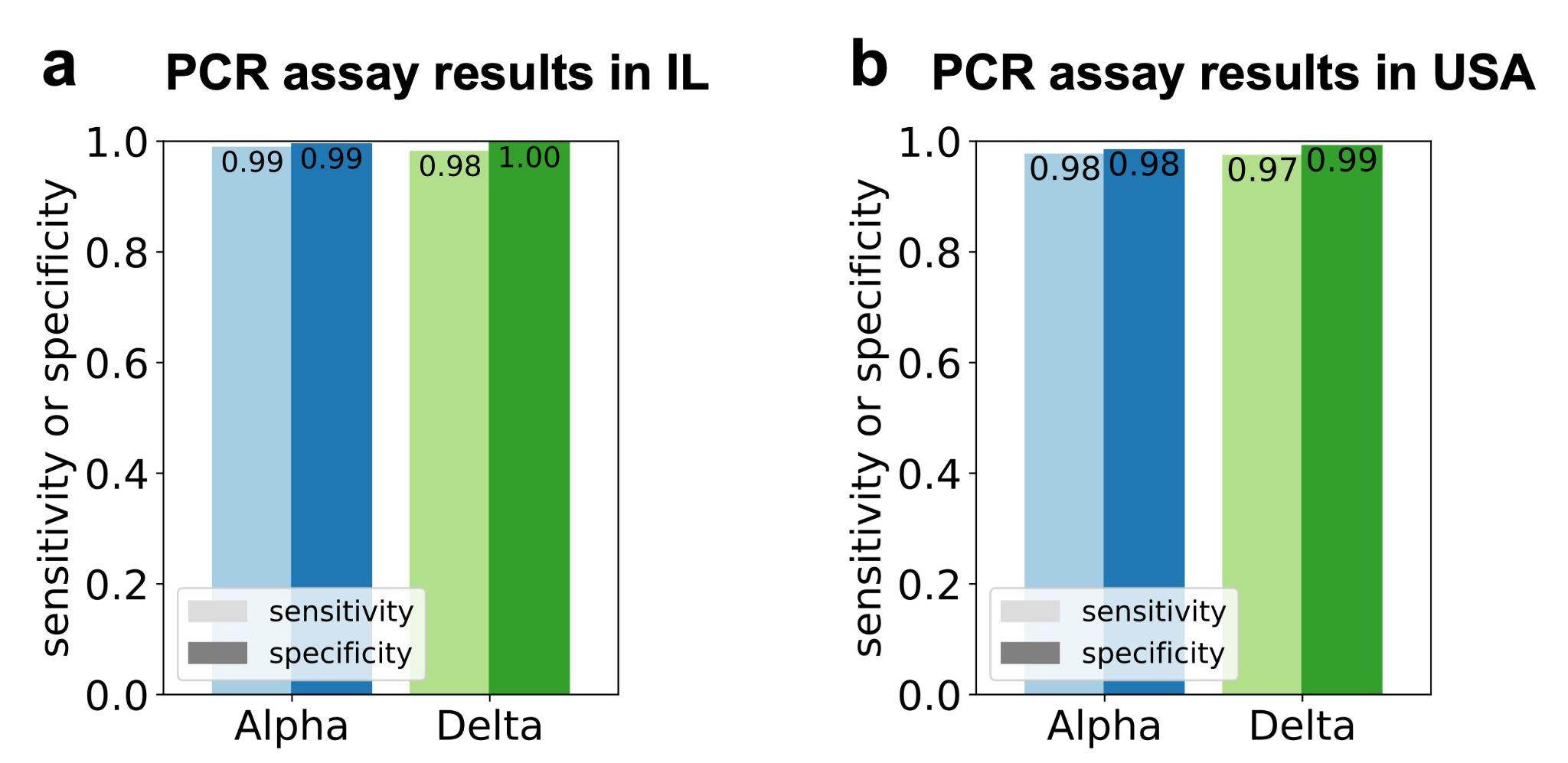
**SI Fig. 4.** Sensitivity and specificity of the two PCR assays designed in this study, one specific to the Alpha variant and the other specific to the Delta variant, estimated using PRIMES in GISAID samples from (a) Illinois and (b) the USA.

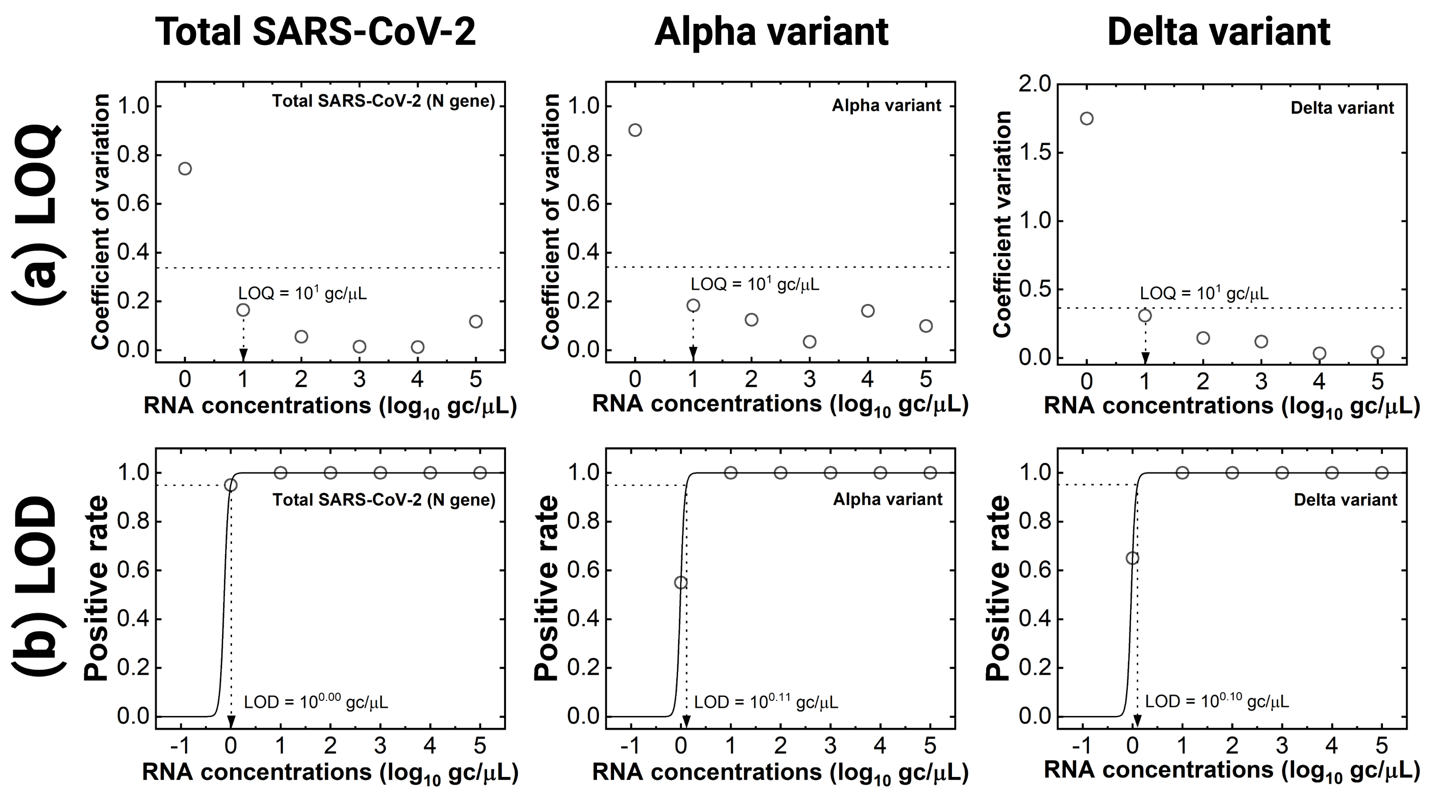

**SI Fig. 5.** Determination of sensitivities (i.e., LOQs and LODs) of RT-qPCR assays for total SARS-CoV-2, Alpha variant, and Delta variant. **(a)** Dashed lines indicate coefficient of variation at 0.35. **(b)** The trendlines for positive rate (solid lines) were calculated by Eq. 6. The LODs were the RNA concentration at which the positive rate is 0.95 (dashed lines).

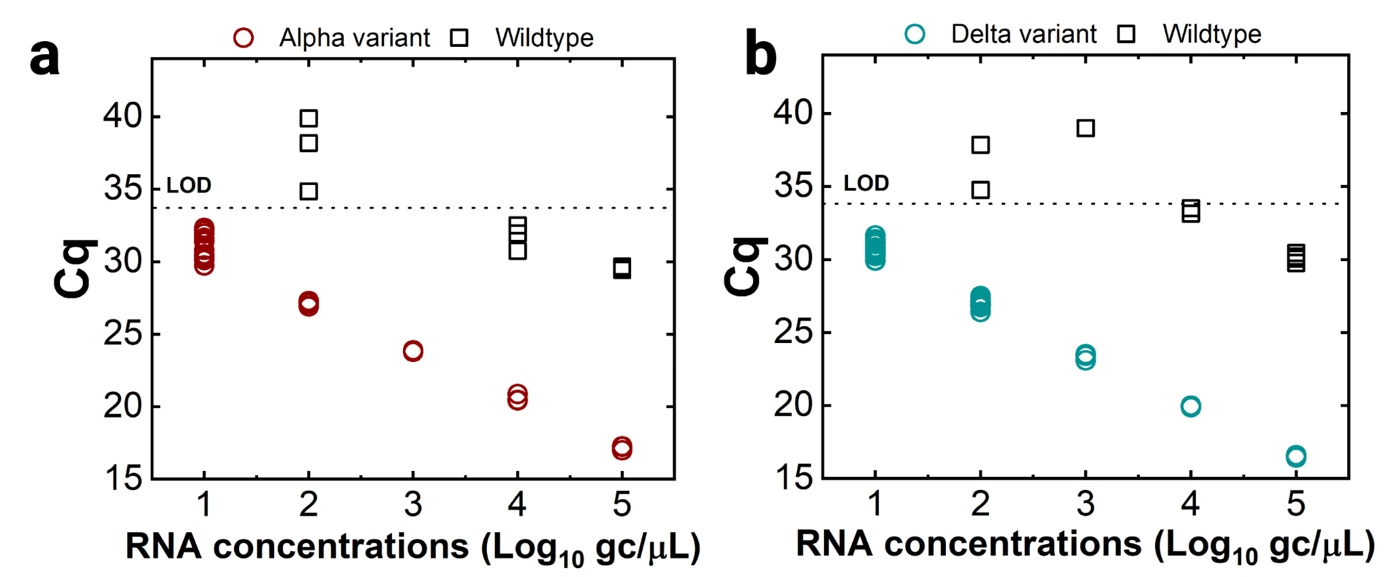

**SI Fig. 6.** Cross-reactivity of PCR assays for Alpha and Delta variants. RT-qPCR assays for **(a)** Alpha and **(b)** Delta variant were applied to its corresponding variant and WT to determine specificity (i.e., cross-reactivity to WT).

**SI Table 3.** Applications of RT-qPCR assays to local sewage samples

| Name | Total SARS-CoV-2 concentration of RNA extracts (gc/µL) | Concentrations (Prevalence) | | Recovery efficiency | Concentration factor | Total SARS-CoV-2 concentration of sewage sample (gc/L) | Decision |
| --- | --- | --- | --- | --- | --- | --- | --- |
|  |  | Alpha variant | Delta variant |  |  |  |  |
| #1 | ${2.7\times10}^{1}$ | ${2.3\times10}^{1}$ (85%) | Below LOD | ${0.58\times10}^{-2}$ | ${0.6\times10}^{-4}$ | ${2.6\times10}^{3}$ | Alpha |
| #2 | ${1.4\times10}^{1}$ | Below LOD | Below LOD | ${0.58\times10}^{-2}$ | ${0.5\times10}^{-4}$ | ${1.3\times10}^{3}$ | Others |
| #3 | ${9.9\times10}^{1}$ | Below LOD | Below LOD | ${0.74\times10}^{-2}$ | ${1.7\times10}^{-4}$ | ${2.3\times10}^{4}$ | Others |
| #4 | ${5.6\times10}^{1}$ | Below LOD | ${3.4\times10}^{0}$ (6%)^1)^ | ${2.37\times10}^{-2}$ | ${4.1\times10}^{-4}$ | ${9.6\times10}^{3}$ | Others |
| #5 | ${1.7\times10}^{2}$ | Below LOD | ${1.5\times10}^{2}$ (92%) | ${0.67\times10}^{-2}$ | ${0.9\times10}^{-4}$ | ${4.3\times10}^{4}$ | Delta |
| #6 | ${1.8\times10}^{2}$ | Below LOD | ${1.3\times10}^{2}$ (73%) | ${0.71\times10}^{-2}$ | ${1.2\times10}^{-4}$ | ${6.0\times10}^{4}$ | Delta |

1) Below the LOQ

**SI Table 4.** The checklist from MIQE guidelines and relevant information for this study

| **Item to check** | **Location** |
| --- | --- |
| 1. Experimental design | |
| Definition of experimental and control groups | Materials and Methods |
| Number within each group | Materials and Methods |
| 2. Sample | |
| Description | Materials and Methods |
| Volume/mass of sample processed | Materials and Methods |
| Processing procedure | Materials and Methods |
| Sample storage conditions and duration | Materials and Methods |
| 3. Nucleic acid extraction | |
| Procedure and/or instrumentation | Materials and Methods |
| Name of kit and details of any modifications | Materials and Methods |
| Contamination assessment (DNA or RNA) | Materials and Methods |
| Nucleic acid quantification | Materials and Methods |
| Instrument and method | Materials and Methods |
| Inhibition testing (C_q_ dilutions, spike, or other) | Materials and Methods |
| 4. Reverse transcription | |
| Complete reaction conditions | Materials and Methods |
| Amount of RNA and reaction volume | Materials and Methods |
| Reverse transcriptase and concentration | Materials and Methods |
| Temperature and time | Materials and Methods |
| Manufacturer of reagents and catalogue numbers | Materials and Methods |
| 5. qPCR target information | |
| Gene symbol | Materials and Methods, Table 3 |
| Sequence accession number | Materials and Methods, Table 3 |
| Location of amplicon | Materials and Methods, Table 3 |
| Amplicon length | Materials and Methods, Table 3 |
| In silico specificity screen (BLAST, and so on) | Materials and Methods, Table 3 |
| 6. qPCR oligonucleotides | |
| Primer sequences | Materials and Methods, Table 3 |
| Manufacturer of oligonucleotides | Materials and Methods, Table 3 |
| 7. qPCR protocol | |
| Complete reaction conditions | Materials and Methods |
| Reaction volume and amount of DNA | Materials and Methods |
| Primer (probe) concentrations | Materials and Methods |
| Polymerase identity and concentration | Materials and Methods |
| Buffer/kit identity and manufacturer | Materials and Methods |
| Manufacturer of plates/tubes and catalog number | Materials and Methods |
| Complete thermocycling parameters | Materials and Methods |
| Manufacturer of qPCR instrument | Materials and Methods |
| 8. qPCR validation | |
| Specificity (gel, sequence, melt, or digest) | Materials and Methods |
| For SYBR Green I, C_q_ of the NTC | Materials and Methods |
| Calibration curves with slope and *y* intercept | Materials and Methods, Fig. 5 |
| PCR efficiency calculated from slope | Materials and Methods |
| *r*2 of calibration curve | Materials and Methods |
| Linear dynamic range | Materials and Methods, Fig. 5 |
| C_q_ variation at LOD | Materials and Methods, SI Fig. 1 |
| Evidence for LOD | Materials and Methods, SI Fig. 1 |
| 9. Data analysis | |
| qPCR analysis program (source, version) | Materials and Methods |
| Method of C_q_ determination | Materials and Methods |
| Outlier identification and disposition | Materials and Methods |
| Results for NTCs | Materials and Methods |
| Description of normalization method | Materials and Methods |
| Number and concordance of biological replicates | Materials and Methods |
| Number and stage of technical replicates | Materials and Methods |
| Repeatability (intra assay variation) | Materials and Methods |
| Statistical methods for results significance | Each figure |
| Software (source, version) | Materials and Methods |
| Data transparency | Raw data available upon request |

**SI Table 5.** The minimum recommended meta-information on sewage samples (2)

| Contents | #1 | #2 | #3 | #4 | #5 | #6 |
| --- | --- | --- | --- | --- | --- | --- |
| Sample location type | Street line manhole | | | | | |
| Population served | ~1700 | ~1700 | ~2400 | ~2160 | ~2160 | ~1100 |
| Combined or separated system | Separated | Separated | Separated | Separated | Separated | Separated |
| Sample collection type | Time-weighted composite samples | | | | | |
| Sample matrix | Sludge from raw wastewater | | | | | |
| Sample date | 01/25/2021 | 02/01/2021 | 01/22/2021 | 02/03/2021 | 09/19/2021 | 09/19/2021 |
| Sample time | ~11 AM | ~11 AM | ~10 AM | ~10 AM | ~10 AM | ~10 AM |
| Sample location | Campus town | Campus town | NE Rantoul | SE Rantoul | SE Rantoul | South Rantoul |
| Pre-concentration storage temperature | Samples were delivered to the laboratory on ice and processed without freezing on the same day. | | | | | |
| Concentration method and citation | RNA extraction from sludge (3) | | | | | |
| Recovery control name & efficiency | BCoV (0.58%) | BCoV (0.58%) | BCoV (0.74%) | BCoV (2.37%) | BCoV (0.67%) | BCoV (0.71%) |
| Extraction method & citation | Viral RNA Mini Kit (Quigen, German) | | | | | |
| Amount of sample processed | 1785 mL | 1855 mL | 585 mL | 245 mL | 1155 mL | 862 mL |
| Extraction blanks results | Signal not detected | | | | | |
| PCR type | qPCR | | | | | |
| SARS-CoV-2 concentrations (gc/L) | ${2.6\times10}^{3}$ | ${1.3\times10}^{3}$ | ${2.3\times10}^{4}$ | ${9.6\times10}^{3}$ | ${4.3\times10}^{4}$ | ${6.0\times10}^{4}$ |
| Target gene | N1 gene (CDC) | | | | | |
| Endogenous wastewater control name & concentration | PMMoV | | | | | |
| Required MIQE guideline | Summarized in SI Table 3 | | | | | |

**SI Table 6.** Information for RT-qPCR assays applied for the sewage samples

| Target species | Target gene or mutation | Primer name | Sequence (5’-3’) | GC content (%) | Tm (℃) | Amplicon size in base-pair (location) | Purpose |
| --- | --- | --- | --- | --- | --- | --- | --- |
| SARS-CoV-2 | N^1)^ | CDC_N1_Forward | GACCCCAAAATCAGCGAAAT | 45.0 | 61.1 | 73  (28287-28358) | Total SARS-CoV-2 |
|  |  | CDC_N1_Reverse | TCTGGTTACTGCCAGTTGAATCTG | 45.8 | 64.5 |  |  |
|  |  | CDC_N1_Probe | ACCCCGCATTACGTTTGGTGGACC | 58.3 | 70.3 |  |  |
|  | S:A570D^2)^ | Alpha_Forward | ACAATTTGGCAGAGACATCGA | 42.9 | 62.3 | 85  (23251-23335) | Alpha variant |
|  |  | Alpha_Reverse | AGAACATGGTGTAATGTCAAGAATC | 36.0 | 61.7 |  |  |
|  | S:P681R | Delta_Forward | ATCAGACTCAGACTAATTCACG | 40.9 | 59.6 | 87  (23583-23669) | Delta variant |
|  |  | Delta_Reverse | TTTCTGCACCAAGTGACATA | 40.0 | 59.7 |  |  |
| PMMOV^3)^ | | PMMOV_Forward | GAGTGGTTTGACCTTAACGTTTGA | 41.7 | 63.4 | 68  (1878-1945) | Normalization to feces |
|  |  | PMMOV_Reverse | TTGTCGGTTGCAATGCAAGT | 45.0 | 63.6 |  |  |
| BCoV^4)^ | | BCoV_Forward | CTAGTAACCAGGCTGATGTCAATACC | 46.2 | 64.2 | 88  (29799-29886) | Recovery efficiency |
|  |  | BCoV_Reverse | GGCGGAAACCTAGTCGGAATA | 52.4 | 63.5 |  |  |
| TV^5)^ | | TV_Forward | GTGCGCATCCTTGAGACAAT | 50.0 | 63.0 | 133  (879-1011) | Inhibition test |
|  |  | TV_Reverse | TTGGAGCCGGGTAGAAACAT | 50.0 | 63.5 |  |  |

1) Taqman-based RT-qPCR was used.
2) [Lee et al., 2021](https://pubs.acs.org/doi/pdf/10.1021/acs.estlett.1c00375) (1)
3) [Haramoto et al., 2013](https://journals.asm.org/doi/full/10.1128/AEM.02354-13) (4); coding sequences for replicase protein were targeted (Genbank: MN496154.1).
4) [Cho et al., 2013](https://www.sciencedirect.com/science/article/pii/S0378113513003337) (5); N gene is targeted (Genbank: LC494177.1).
5) Fuzawa et al., 2020 (6); FLA45_gp1 gene is targeted (Genbank: NC_043512.1).
6) Primer pair specificity was checked by the primer-blast tool (National Center of Biotechnology Information). Each pair of primers were blasted with Homo sapiens (txid:9606). We confirmed that our primers do not target any sequences of their host cells.
